# The impact of quality control on cortical morphometry comparisons in autism

**DOI:** 10.1101/2022.12.05.22283091

**Authors:** Saashi A. Bedford, Alfredo Ortiz-Rosa, Jenna M. Schabdach, Manuela Costantino, Stephanie Tullo, Tom Piercy, Lifespan Brain Chart Consortium, Meng-Chuan Lai, Michael V. Lombardo, Adriana Di Martino, Gabriel A. Devenyi, M. Mallar Chakravarty, Aaron F. Alexander-Bloch, Jakob Seidlitz, Simon Baron-Cohen, Richard A.I. Bethlehem

## Abstract

Structural magnetic resonance imaging (MRI) quality is known to impact and bias neuroanatomical estimates and downstream analysis, including case-control comparisons. However, despite this, limited work has systematically evaluated the impact of image and image-processing quality on these measures, or compared different quality control (QC) methods and metrics. The growing size of typical neuroimaging datasets presents an additional challenge to QC, which is typically extremely time and labour intensive. Two of the most important aspects of MRI quality are motion, which is known to have a substantial impact on cortical measures in particular, and the accuracy of processed outputs, which have been shown to impact neurodevelopmental trajectories. Here, we present a tool, FSQC, that enables quick and efficient yet thorough assessment of both of these aspects in outputs of the FreeSurfer processing pipeline. We validate our method against other existing QC metrics, including the automated FreeSurfer Euler number, and two other manual ratings of raw image quality. We show strikingly similar spatial patterns in the relationship between each QC measure and cortical thickness; relationships for cortical volume and surface area are largely consistent across metrics, though with some notable differences. We next demonstrate that thresholding by QC score attenuates but does eliminate the impact of quality on cortical estimates. Finally, we explore different ways of controlling for quality when examining differences between autistic individuals and neurotypical controls in the ABIDE dataset, demonstrating that inadequate control for quality can alter results of case-control comparisons.

## Introduction

It is well established that magnetic resonance imaging (MRI) quality affects neuroimaging-derived neuroanatomical measures ^1^. Of particular concern is in-scanner head motion, which has been consistently shown to affect estimates of brain structure ^1–7^, function ^8–11^ and connectivity ^12,13^. While functional magnetic resonance imaging (fMRI) studies were initially the primary focus of such research ^8–10^, more attention has recently been brought to the impact of motion on structural MRI studies. For example, estimates of cortical thickness, surface area, and volume have consistent, regionally dependent relationships with motion ^1,2,4–6^. In addition to motion, other factors such as scanning artefacts, intensity inhomogeneities, and geometric and susceptibility-related distortions also impact image quality ^6^. Critically, image quality, and head motion in particular, is highly correlated with demographic characteristics such as age, sex, as well as variables of interest such diagnostic status in clinical cohorts ^4–6^. Errors in image processing outputs and overall image quality also significantly impact and distort estimates of neurodevelopmental trajectories especially ^6,14,15^, and there is evidence that these biases also permeate case-control comparisons ^16,17^. Although these issues are becoming more widely acknowledged, there is currently no “gold standard” of quality control (QC) methods. Detailed QC procedures are also rarely reported, making quantitative evaluations across studies difficult. Here we sought to quantitatively evaluate several QC procedures and their association with resulting morphometric measures, as well as quantitatively assess the impact of QC procedures on case-control differences in the context of autism.

One barrier to implementing thorough and rigorous QC is the increasing sample sizes typically used in neuroimaging studies ^18–24^, in particular when using publicly available datasets. Manual QC is both time and labour intensive, and requires expert raters and/or extensive training of individuals to examine and assess both raw scans and post-processed outputs, as well as assessment of inter-rater reliability ^15,16,25^. With samples routinely in the thousands or even tens of thousands, this is often impractical or infeasible. In recent years, various alternative, automated QC methods have been proposed. For example, FreeSurfer’s Euler number is a good proxy for image quality, correlating highly with manual quality ratings, as well as regional measures of cortical thickness ^15^. Other recently developed approaches, such as MRIQC ^26^ and Qoala-T ^27^ provide automated reports of image quality, and prediction of manual quality ratings, based on various image quality metrics, which include measures of noise, entropy (indicative of motion), statistical properties, cortical features and extreme values, and specific artefacts. Another approach is to use “citizen science”, combined with manual expert ratings and machine learning, to generate thousands of QC ratings, lessening the burden on researchers. This approach has resulted in the Swipes for Science initiative (swipesforscience.org), which crowd-sources QC ratings (binary pass/fail classification) of raw images, and also accounts for variations in quality of ratings by different raters ^28^.

Few extensive and detailed manual quality control protocols have been explicitly published ^29^. While authors sometimes summarise QC procedures in methods sections or supplementary results ^4,16^, more commonly little detail is given. Some papers have provided and assessed detailed protocols for QC of post-processing outputs. For example, Visual QC ^30^ and a QC protocol provided by the ENIGMA consortium ^31^ provide detailed guidelines and a framework in which to view and rate images and their FreeSurfer outputs. While these protocols offer a comprehensive and useful tool for evaluating scans and surface reconstructions, they are time consuming, hence may be impractical for very large datasets. It also remains an open question whether manual QC procedures outperform automated methods and thus warrant the extensive time and effort required. The lack of consensus or standardised methods is particularly problematic for large publicly available datasets, as it makes comparisons between different studies using the same datasets challenging and it is unclear to what extent inconsistencies in results are due to inconsistent QC methods or standards.

This is a particularly salient issue in neurodevelopmental imaging, as inadequate image quality has been shown to impact findings ^6,14^, and participants with neurodevelopmental conditions such as autism are more susceptible to image quality issues (often due to motion) than neurotypical individuals ^4,5,16^. Without adequate QC, there is a high risk of spurious correlations or group differences, as well as true effects being obscured by motion or quality issues. Numerous studies have used the Autism Brain Imaging Data Exchange (ABIDE) ^21,32^ to examine case-control differences related to autism, using both structural and functional measures ^16,33–44^. Although there is some convergence of these findings, there are also conflicting and inconsistent findings between studies, which may in part be due to differences in QC procedures and thus differences in the final sample. The issue of how extensively variations in quality impact neuroanatomical estimates urgently warrants further investigation, in particular in relation to neurodevelopmental and psychiatric conditions.

Given the need for systematic, rigorous, and reproducible QC methods, we aimed to develop an efficient yet thorough tool for QC of FreeSurfer surface reconstructions that also captures aspects of raw image quality, specifically motion. We also aimed to validate our QC metric against other manual and automated QC methods in the ABIDE dataset. Finally, we assessed the impact of image quality on regional estimates of cortical morphometry, and examined the interaction between image quality and diagnostic status in the context of autism.

## Methods

### Sample

The ABIDE dataset consists of neuroimaging, demographic, and clinical data from 2226 individuals (1060 autistic individuals and 1166 neurotypical controls), aged 5-64 years (1804 assigned-males-at-birth, 422 assigned-females-at-birth). The ABIDE repository includes two waves of data aggregation (ABIDE I and II), from a total of 24 international sites. Participant demographics and acquisition information have been previously described in detail ^21,32^.

### FreeSurfer QC method and generation of images

#### Processing with FreeSurfer

All T1-weighted structural scans were processed with FreeSurfer 6.0.1 (see ^21,32^ for details on ABIDE acquisition). Cortical parcellations were derived using both the Glasser ^45^ and Desikan-Killianey ^46^ atlases.

#### Generation of FSQC images

QC images were generated by overlaying the FreeSurfer-derived cortical surface boundaries on the participant’s T1 scan in FreeSurfer’s FreeView visualisation tool, and using the FreeView Screenshot function to generate screen captures at different views and slices of the brain (3 axial; 3 coronal; 4 sagittal, for a total of 10 images per subject; see Figure 1). This process was automated in a virtual server window, with consistent coordinates specified for each participant for the 10 screenshots (code shared below).

**Figure 1.**
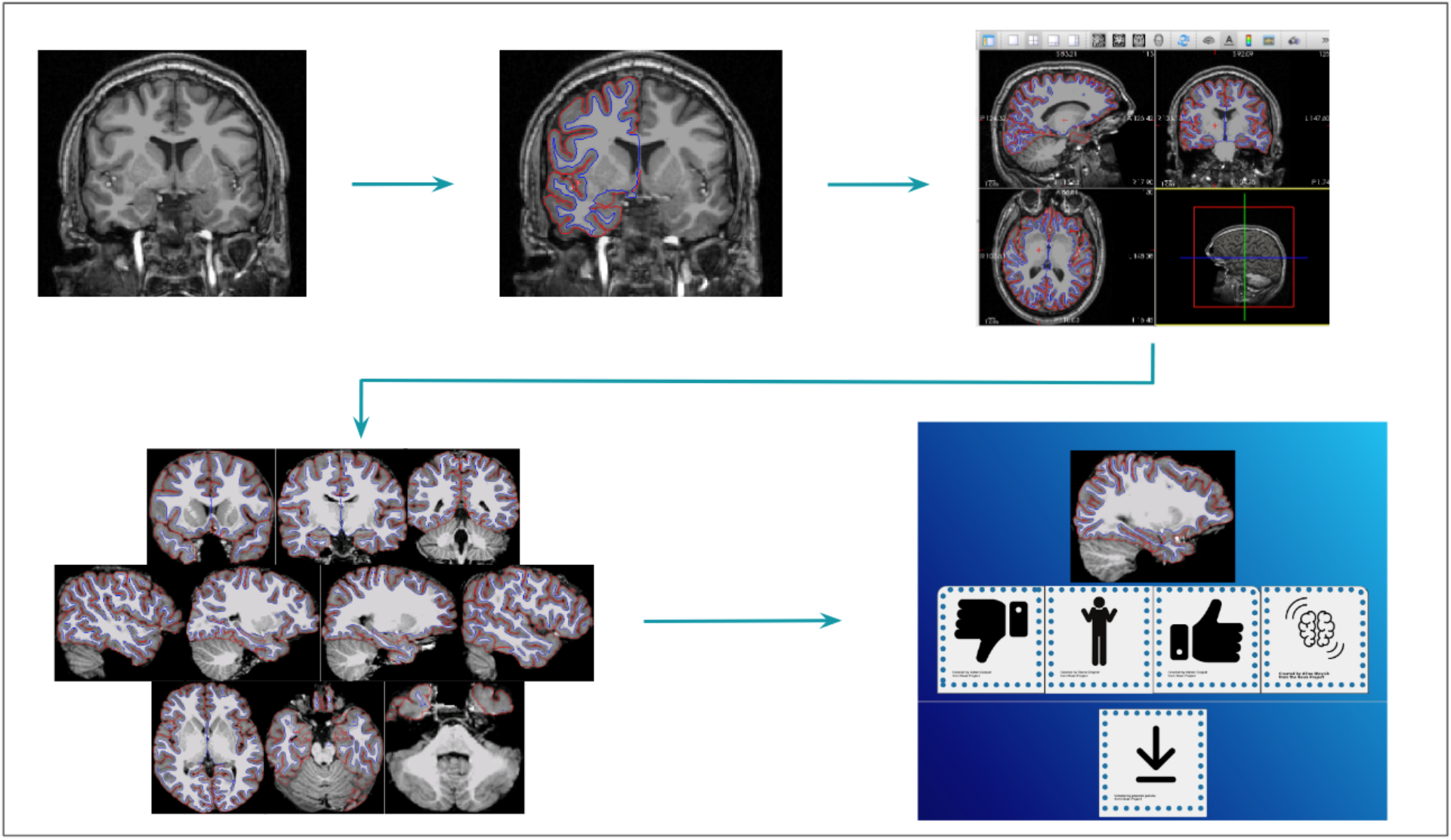
FSQC image generation workflow. From left to right: T1 images were processed with FreeSurfer 6.0.1 and displayed in FreeView with pial and white matter surfaces overlaid on the T1 image (both hemispheres). Screenshots were automated and taken at predefined, consistent coordinates, for a total of 10 images per participant. Images were then displayed and rated in the Image-Rating app, and scores were averaged across all 10 images for each participant.

Prior to rating, each image was renamed using the MD5 message digest algorithm and images were randomly shuffled so ratings were not biased by other images from the same participant appearing in sequence. Images were then viewed in the Image-Rating QC application (https://github.com/sbedford0/FSQC/imageratingQCApp), and assessed for accuracy of the cortical reconstruction (grey-white matter and grey matter-pial surface boundaries), as well as presence of motion in the raw T1 image on which the surfaces were overlaid. Each image (10 per participant) was rated individually on a scale of 1 - 4 (good - bad), corresponding to the following categories: good (1), minor error (2; i.e., often involving misestimation of boundaries restricted to one or two specific regions), visible motion (3; e.g., ringing, blurring of scan), and bad (4). See https://dx.doi.org/10.17504/protocols.io.kxygx9m6wg8j/v1 for images, detailed criteria and examples of each score. Outputs from the Image-Rating app were recorded and downloaded in a csv file. Categorical ratings were then converted to the corresponding numerical rating (i.e., 1-4), and averaged across all 10 images for each participant, to give a final continuous score between 1 and 4 per participant. Thus, these scores provide a quality rating reflecting the accuracy of the FreeSurfer surface reconstructions as well as motion in the raw T1 image.

### Statistical analysis

#### FSQC inter-rater reliability

Two raters (SB and RB) rated the entire dataset, and an average of the two scores was taken for each participant. To ensure reasonable inter-rater reliability, raters first rated a subset of ∼200 images, which were compared, and any major discrepancies or issues resolved, before moving on to the rest of the dataset. This consensus rating was also used to clarify any discrepancies in the image rating protocol. To assess inter-rater reliability of the method and protocol across multiple raters, 6 raters (SB, RB, AOR, JS, AAB, JMS) assessed a subset of 50 participants (500 images); Spearman’s correlations and two-way ICC for agreement were calculated across all raters, on the average score for each participant.

The first main analysis (examining the effect of FSQC on cortical thickness, see below) compared both individual rater’s scores, as well as the average score across all raters to ensure consistency (supplementary methods 1.1). For all subsequent analyses using FSQC, the average scores between the two raters was used.

#### Relationship between different QC metrics

First, we sought to validate our FSQC method by examining the relationship between FSQC scores (averaged across 10 images per participant, and two raters), and other QC metrics. These included the FreeSurfer-derived Euler number ^15^; a manual score assessing the presence and amount of motion in each image (“Motion QC”; raters SB, MMC, ST ^16^, see https://github.com/CoBrALab/documentation/wiki/Motion-Quality-Control-%28QC%29-Manual); and another manual rating of overall image quality which was derived from and built upon “Motion QC” (“PondrAI QC”; raters MC, GAD). Spearman correlations were run to assess the relationship between FSQC and each other metric.

#### Demographic correlations

Since demographic factors are related to image quality ^4,5,16^, we next investigated these relationships in our dataset. Of particular interest were age, sex-assigned-at-birth (hereafter “sex”), and diagnosis, as these variables are especially relevant to neuroimaging studies of autism and have been shown by previous work to correlate with image quality, and motion specifically ^2,4,5^. Linear mixed effects models were used to examine the impact of age, a quadratic term for age (age^2^), sex and diagnosis (with site as a random effect) on all four quality metrics separately (FSQC, Euler, Motion QC, PondrAI QC).

#### Impact of QC on cortical morphometry

To examine and quantify the impact of image quality, as measured by all QC metrics, on different neuroanatomical measurements, we assessed the relationship between each QC measures and global neuroanatomical measures, including total cortical and subcortical grey matter volumes (cGMV and sGMV), total brain volume (TBV), total white matter volume (WMV), total ventricular volume, and mean cortical thickness.

As previous work has demonstrated spatially dependent relationships with quality ^2,4,5^, we next examined regional effects on cortical thickness (CT), surface area (SA) and cortical volume (CV), using Glasser parcellations. Relationships with subcortical phenotypes were not assessed as the surface reconstructions being rated in the FSQC tool include only the cortical surface boundaries. Analyses were initially run on all participants (i.e., no exclusions), to examine the relationship between different types of quality and cortical morphometry across the whole spectrum of quality. For these analyses, linear mixed effects models were run for each parcellation across the brain, separately for CT, SA and CV. All regression models included QC metric, age, age^2^, and sex as fixed effects, and site as a random effect, with CT/SA/CV as the dependent variable, for each region. Partial correlations were calculated to quantify the strength of the association between QC metric and neuroanatomical measure (e.g., FSQC and CT; motion QC and CV, etc). Results were corrected for multiple comparisons using the false discovery rate (FDR) across parcellations in all analyses. For subsequent analyses, we focus on FSQC, our newly developed quality metric, and Euler, a commonly used automated method.

Supplementary analyses were also run using Desikan-Killianey parcellations, for comparison with previous work (supplementary methods 2.1). The main analyses were repeated using a random effects meta-analysis for comparison, and to assess heterogeneity of results across sites (supplementary methods 2.2). We also attempted to replicate these analyses in a larger, more representative dataset of 74,647 individuals (that has been previously used^18^; supplementary methods 2.3). Finally, we conducted a variance partitioning analysis to evaluate the relative contribution of image quality to the total variance explained, compared to factors such as diagnosis, age, sex and site (supplementary methods 2.4).

#### Exclusion/thresholding analyses

Quality control scores are often used as a way to exclude data of poor quality; for example, previous studies using the Euler number as a QC metric recommend a study-specific threshold ^15^. To evaluate the impact of different quality thresholds on the relationship with cortical morphology and existence of group-level differences, and to assess the extent to which results were driven by participants with the worst or more extreme image quality, we conducted a thresholding analysis, examining the impact of quality (FSQC and Euler number) at cut-offs of varying stringency.

First, for FSQC, we chose score thresholds in increments of 0.5 points (3, 2.5, 2, 1.5). The same models and analyses described above were re-run after excluding participants at each of these thresholds, for each cortical phenotype. For Euler number, as there were less obvious cut-off points than for FSQC, we used median absolute deviations (MAD) to determine various thresholds for these analyses. The relationship between Euler and each cortical phenotype were assessed after thresholding at 1, 2 and 3 MADs, and half points in between (corresponding to Euler numbers of 139, 174, 210, 245, 281 and 317). Due to the significant differences and variability between sites, supplementary analyses were also conducted applying MAD-based cut-off points calculated and applied individually per site, rather than across the whole sample (supplementary methods 3.1).

Additional sensitivity analyses were performed, including comparing high and low quality based on a median FSQC split, and thresholding based on the top percentage of scores (applied to the whole sample and per site) (supplementary methods 3.2-3.3).

#### Interaction between image quality and diagnosis

As image quality differs by diagnostic status and impacts neuroanatomical estimates ^2,4,5^, it is likely that inadequate accounting for quality will lead to inaccurate conclusions relating to diagnostic differences. To this end, we examined differences in cortical morphometry between autistic individuals and controls with different methods of accounting for quality, and at different quality thresholds. First, we examined group differences in CT, SA and CV without accounting for quality, using linear mixed effects models with diagnosis, age, age^2^ and sex in the model, and site as a random factor. Next, the same models were run with the addition of FSQC or Euler number as a covariate to assess the impact of controlling for quality, as well as thresholding by both FSQC (at 2.5) and Euler (at 2 MAD).

Supplementary analyses for CT examined diagnostic effects after thresholding by FSQC or Euler at various cut-off points (FSQC: 3, 2.5, 2, 1.5; Euler: 1, 2 and 3 MADs; supplementary methods 4.1), as well as the effect of diagnosis on CT after thresholding by FSQC and also controlling for Euler (supplementary methods 4.2). Finally, we examined the interaction between diagnosis and FSQC or Euler on CT (supplementary methods 4.3).

### Data and code availability

The imaging rating tool, code to generate QC png images and analysis scripts are available at: https://github.com/sbedford0/FSQC. The full protocol can be found at: https://dx.doi.org/10.17504/protocols.io.kxygx9m6wg8j/v1.

## Results

### Inter-rater reliability

For the subset of 50 participants, the ICC was moderate, at 0.68 for all 6 raters. Spearman correlations calculated between each pair of raters ranged from 0.68-0.86 (see Figure 2A). For the whole dataset the inter-rater Spearman correlation was 0.63 between raters SB and RB.

**Figure 2.**
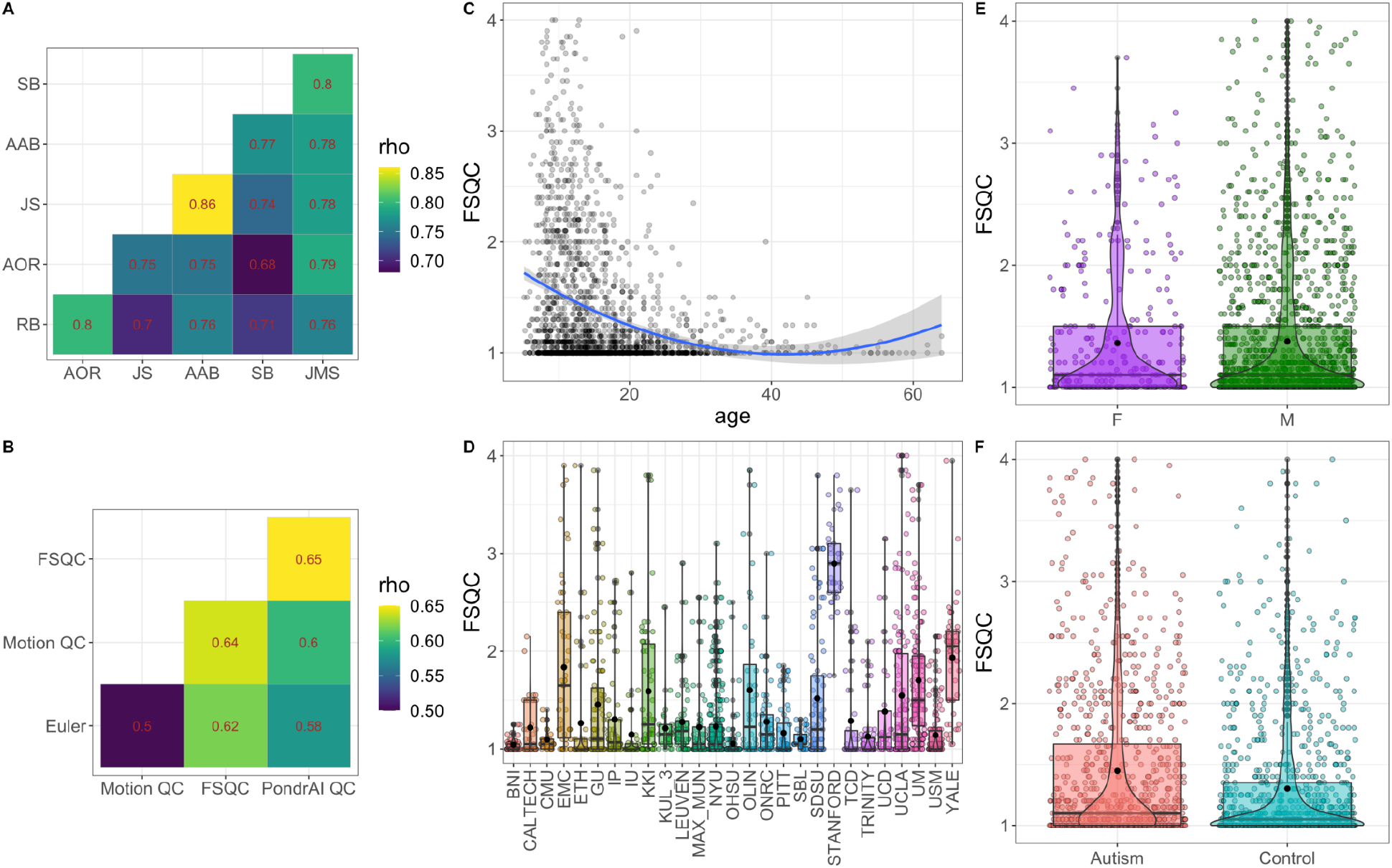
**A**. Inter-rater correlation matrix for FSQC ratings for a subset of 50 participants (500 images). All pairs of raters were significantly correlated with each other between 0.7-0.8 rho. **B**. Correlations between different QC metrics. All measures were significantly positively correlated with each other between 0.5-0.65 rho. **C**. Relationship between FSQC and age. A significant effect of age was observed in which younger participants had lower quality ratings. **D**. FSQC score distributions by site. There was significant variability in quality across sites. **E**. FSQC distributions for males and females. There was no significant sex difference in FSQC. **F**. FSQC distributions by diagnosis. Autistic participants had significantly higher FSQC scores (i.e. lower image quality) relative to controls (p<0.0001, *d* = -0.27).

Results of the impact of FSQC on CT were nearly identical when using each rater’s scores separately (SB and RB), and the average of the two scores (see supplementary figure S1.1).

### Relationship between different QC metrics

FSQC was significantly and positively correlated with all other measures (Euler number (rho=0.62, p<0.0001); Motion QC (rho=0.64, p<0.0001); PondrAI QC (rho=0.65, p<0.0001) (Figure 2B).

### Demographic correlations

We assessed the relationship between each metric and demographic variables previously reported to be highly correlated with image quality (Figure 2C-F). For all quality measures, autistic participants had significantly lower image quality relative to controls (all p<0.01; Cohen’s *d* = -0.14 - -0.29). For all metrics, there was also a significant effect of age and age^2^ (p<0.0001). However, when we examined the relationship between age and quality in young and old groups after performing a median split, both groups showed a negative relationship, reflecting lower image quality in younger participants. For motion QC only, there was a significant effect of sex, where males had significantly worse quality scans than females (p=0.004, Cohen’s *d* = 0.16). Image quality, across all metrics, also differed significantly by site (p<0.0001).

### Impact of QC on cortical morphometry

FSQC was significantly but weakly correlated with global brain measures of total cortical GMV, WMV, subcortical GMV and TBV at a Bonferroni-corrected threshold of p<0.008 for six comparisons (rho = -0.07 - -0.16), but not with mean CT or ventricular volume (see supplementary table S1 for all correlations). Regional analyses revealed significant associations across much of the cortex for all cortical phenotypes and QC metrics, passing 5% FDR (partial r = -0.46-0.40). Associations were largely negative, denoting apparent decreases in cortical measures with lower quality (higher scores), though strong positive relationships (increased measures with lower quality) were observed in some regions. Each phenotype showed distinct spatial relationships with quality; however spatial patterning across the cortex was strikingly similar between all metrics within each phenotype, with slight differences observed only in the case of Euler number for SA and CV. Results of regional analyses are shown in Figure 3, showing partial r values thresholded for significant regions (surviving 5% FDR). For maps of all (including non-significant) partial r values across the cortex, see supplementary figure S2.

**Figure 3.**
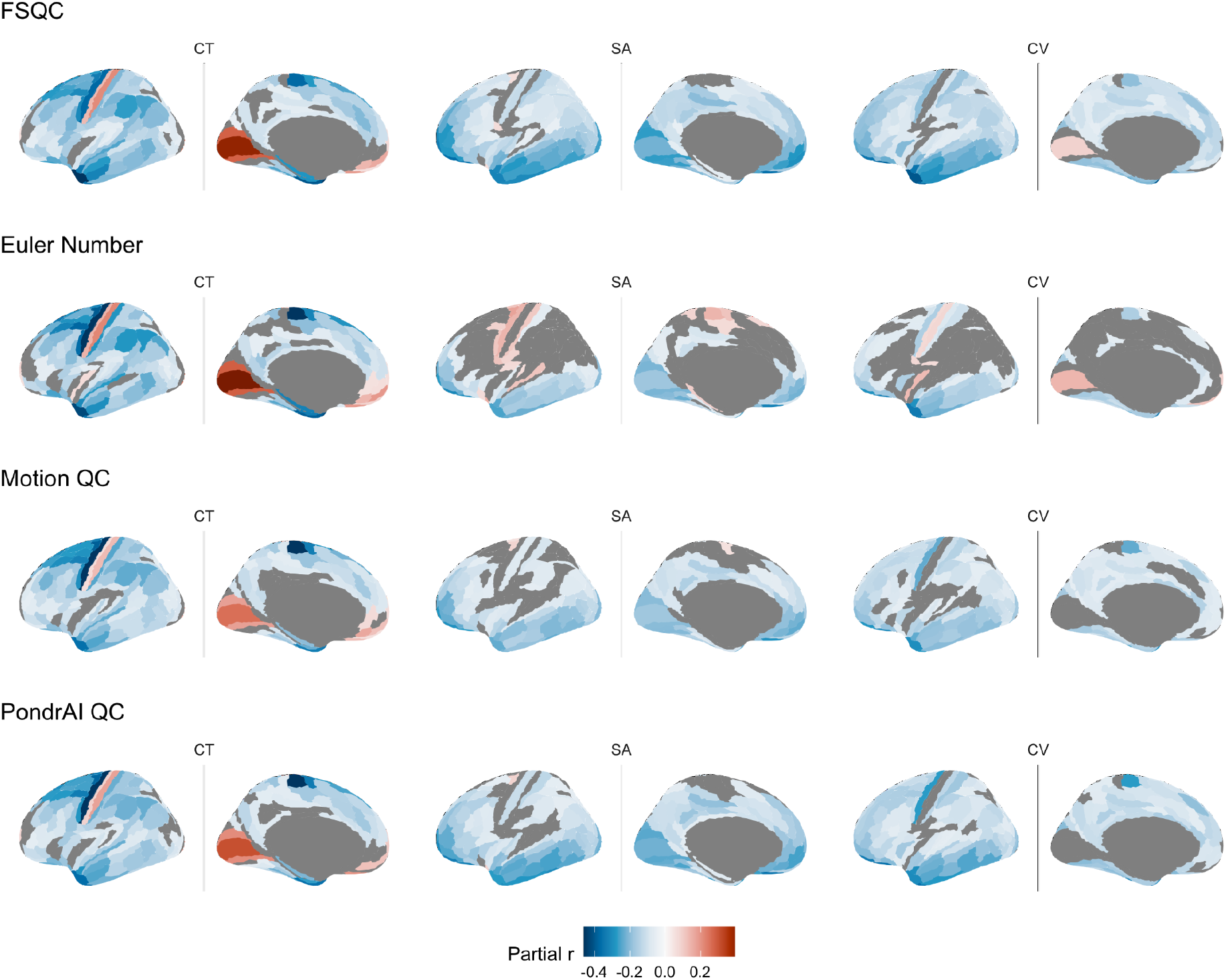
Associations between QC metrics and regional cortical morphometry. There was a significant relationship between image quality and neuroanatomical estimates across much of the cortex for all metrics and phenotypes. Relationships were largely negative, and were strongest for cortical thickness. Spatial patterning of results was highly similar across metrics, with the exception of Euler number of SA and CV, which showed more positive associations, but less significant relationships overall, than the other three measures.

Of the three cortical phenotypes, the strongest associations were observed for CT, and as such were the main focus of subsequent analyses (with CV and SA results reported in supplementary materials). The strongest negative correlations between CT and image quality (across metrics) were observed in lateral superior frontal (including precentral gyrus), parietal, and inferior temporal regions, with widespread weaker, but still significant, negative correlations across much of the frontal, parietal and temporal cortices. Significant positive correlations were observed in the medial occipital and ventromedial prefrontal cortices for all metrics, as well as in the postcentral gyrus (Figure 3).

The strongest significant negative correlations for surface area were observed in inferior (medial and lateral) frontal and temporal cortices, as well as the medial occipital cortex. Correlations and spatial patterning were again mostly consistent across QC metric, with the exception of a larger number of positive correlations, and slightly fewer significant correlations overall, observed for Euler number. For Motion QC and PondrAI QC, almost no positive correlations reached significance, and in FSQC, only two or three disparate regions (including the postcentral gyrus) showed positive significant correlations. For Euler number, by contrast, significant positive correlations were observed in regions including the pre- and postcentral gyrus, medially and laterally, as well as the superior temporal gyrus (Figure 3).

Spatial patterning for cortical volume was again very similar across metrics with the exception of Euler. The medial occipital cortex was significantly positively correlated with QC in FSQC and Euler, but did not reach significance in the other two metrics, though subthreshold correlations were also positive. For all metrics except Euler, significant but weak negative correlations were observed across much of the cortex, and most strongly in inferior temporal and frontal regions, and the precentral gyrus. For Euler, less regions met significance, including large areas of the frontal and parietal cortices. Most significant correlations were still negative for Euler, though positive correlations were observed in the postcentral gyrus, medial prefrontal cortex, and left hippocampus (Figure 3).

Desikan-Killiany parcellations and results of the meta-analytic technique both displayed consistent spatial patterning. Our replication analysis in over 100,000 individuals also yielded largely consistent results (supplementary results S2.1-2.3). The variance partitioning analysis indicated FSQC and Euler contributed a relatively small portion of the variance, but larger than diagnosis (supplementary results S2.4).

Almost all analyses showed the strongest effects for cortical thickness, consistent with previous work suggesting that CT is more susceptible than other cortical estimates to impacts of image quality and motion ^4^. Consequently and for clarity, subsequent analyses will focus primarily on the relationship between CT and image quality. For cortical surface area and volume results, see supplementary results.

### Exclusion/thresholding analyses

We next examined the impact of different levels of QC thresholding stringency on the relationship between quality and cortical morphometry, based on FSQC and Euler number. For CT, after excluding only scans with the worst FSQC scores (3 and above), effect sizes for the association with FSQC were attenuated, but significant associations were still observed across much of the cortex, following the same spatial patterns as the non-thresholded analysis. Effect sizes were further attenuated, but with similar patterning (strongest results retained) after excluding those with scores higher than 2.5. After excluding at scores of 2 and 1.5, few regions maintained significant associations with FSQC (inferior frontal and temporal regions, and superior frontal cortex and precentral gyrus; Figure 4).

**Figure 4.**
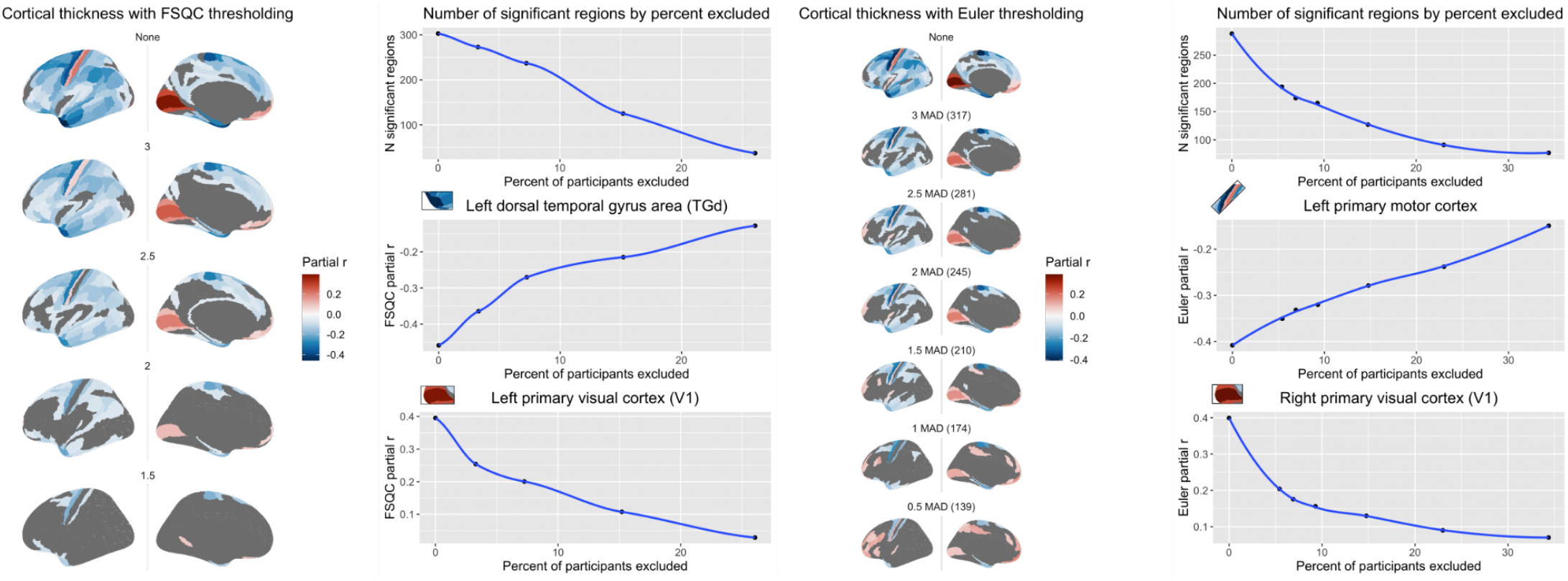
Relationship between cortical thickness and FSQC (left) and Euler number (right) after thresholding at different levels of stringency. Accompanying graphs show the attenuation of both number of significant regions observed (top) and partial correlation effect size (bottom two panels) as stringency increases.

In the Euler MAD-based thresholding analysis, we observed similar but slightly less stark differences between cut-off points than with FSQC. For CT, an attenuation of effect size was still observed, but was somewhat more gradual and to a lesser extent than when using FSQC. The maps for cut-off points of between 2-3 MAD looked similar, with more of a substantial drop off in significant regions after a cut-off of 1 MAD (Figure 4). Additional sensitivity analyses all yielded similar results (Supplementary figures 3.1-3.3).

SA and CV showed a more stark and immediate drop off in significant effects in the FSQC thresholding analyses (supplementary Figure S3.4). Interestingly, in the Euler thresholding analyses for SA and CV, rather than an attenuation of significant effects, we observed a change in direction, such that associations with Euler number went from mostly negative to mostly positive after thresholding (Supplementary figure 3.5).

### Interaction between image quality and diagnosis

There were minimal differences in cortical morphometry between autistic and neurotypical controls when not accounting for image quality. Autistic individuals had greater CT in the medial primary visual cortex (V1), and a small region in the medial parietal lobe relative to controls, and thinner cortex in a few small regions in the left superior frontal and inferior prefrontal cortex. The effect of controlling for FSQC and Euler number were similar. In these analyses, right V1 was no longer significant; nor were any of the regions which had shown thinner cortex in autism, with the exception of the inferior prefrontal cortical region. Additionally, after controlling for either QC metric, additional significant effects (greater CT in autism relative to controls) were observed in the superior temporal gyrus. Though not many regions survived FDR in any analysis, when examining subthreshold results, we noted that most of the effects that were diminished or disappeared after controlling for quality were those in which apparent thinner cortex in autistic individuals was observed in the original analysis, suggesting that these results may have been an artefact of poor image quality (in the autistic group in particular).

Results were similar, although not identical, after applying QC thresholding (for Euler or FSQC) instead of simply controlling for quality. Results were essentially the same whether applying a cut-off based on FSQC or Euler at a similar stringency (FSQC cut off of 2.5 [N = 1727]; Euler threshold of 2 MAD or 245 [N = 1689]): only regions in the bilateral medial occipital cortices, and right medial parietal cortex remained significant, all of which were thicker in autism than controls. Again, all regions with thinner cortex in autism were no longer significant after thresholding (Figure 5). Results did not change substantially when applying thresholds of different levels of stringency based on FSQC or Euler, though were slightly further attenuated at each cut-off point (supplementary results S4.1).

**Figure 5.**
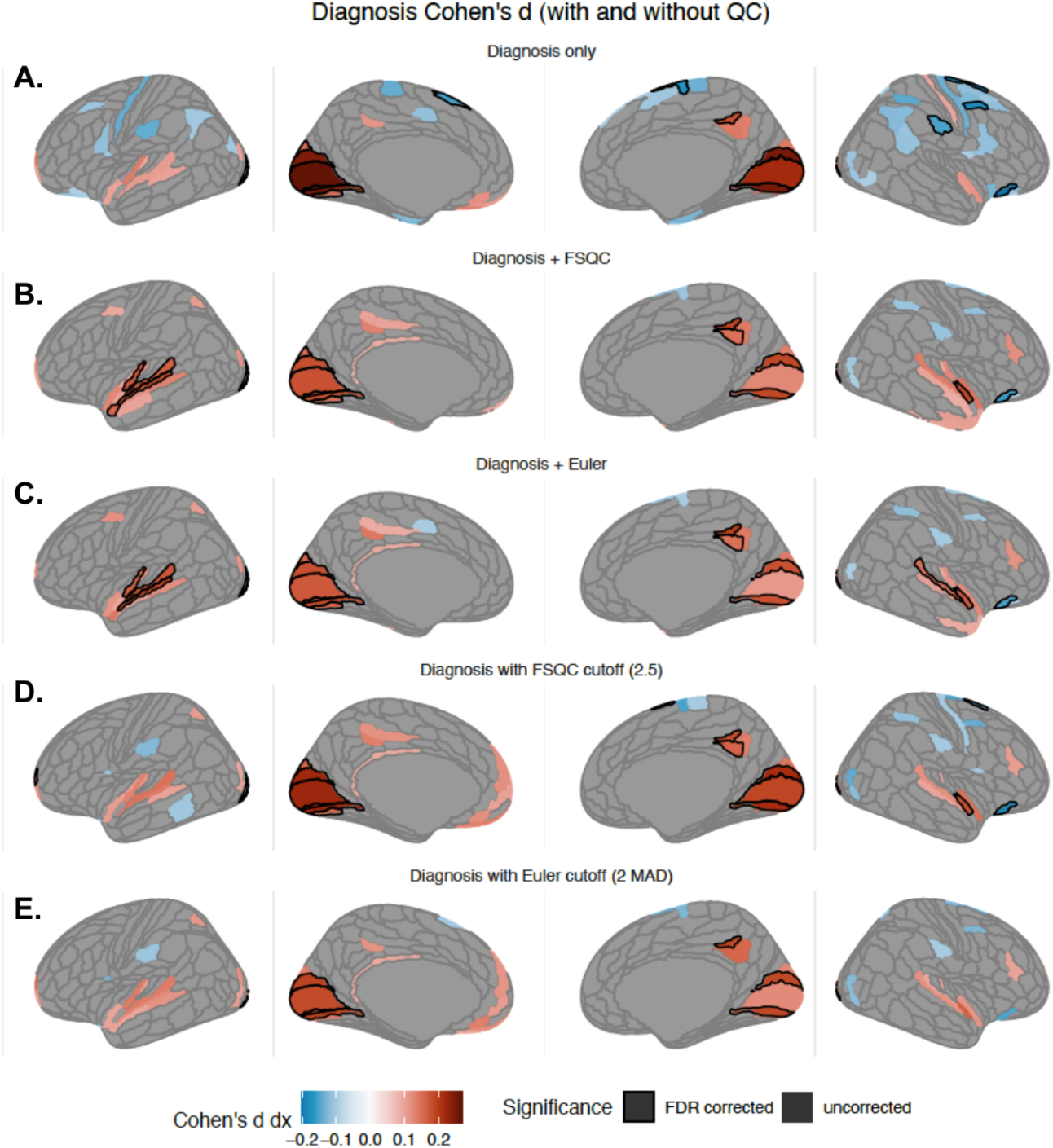
Impact of autism diagnosis on cortical thickness (Cohen’s *d*) without accounting for image quality (A), when controlling for FSQC (B) or Euler (C), and thresholding by FSQC (D) and Euler (E). Significant regions passing 5% FDR are shown with a black border; other regions are subthreshold (i.e., not surviving FDR) differences. Most results indicate thicker cortex in autism relative to controls; results do not change drastically with quality control, but most negative associations between diagnosis and CT (autism < controls) disappear. Significantly thicker cortex in the superior temporal gyrus, which has previously been reported in autism, is observed only when controlling for quality (FSQC or Euler).

Combining the two approaches by applying a threshold based on FSQC while also controlling for Euler did not drastically change the results, though some additional regions showed significant associations (supplementary figure S4.2). The interaction between quality and diagnosis suggested a stronger relationship between quality and cortical thickness in the autistic group than controls (supplementary figure S4.3). Only very minimal group differences in SA and CV were observed, both with and without accounting for image quality (supplementary results S4.4).

## Discussion

Our results demonstrate significant, widespread associations between image quality and cortical morphometry across the brain, which are largely consistent across multiple QC metrics. These QC-morphometry interactions persist even after excluding participants with lower image quality, and have profound effects on case-control evaluations. We have outlined several ways to evaluate and correct for the issue of image quality and empirically show that these can improve the consistency and robustness of clinical neuroimaging findings.

### The FSQC tool enables fast and robust evaluation of image quality in a scalable manner

Our FSQC tool is easy and quick to implement even for large datasets, while still being rigorous and thorough. The generation of multiple images per participant, at multiple orientations and slices across the cortex, allows for a thorough examination of different views without the time consuming process of individually opening and scrolling through each scan slice by slice. Importantly, it takes into account both raw image quality (e.g., motion), and quality of FreeSurfer post-processing outputs and surface reconstructions, simplifying the QC process. Finally, we have shared both our FSQC tool and protocol, and completed image ratings for ABIDE, with the neuroscience community. This could help to save other researchers unnecessary time and effort, and help to improve consistency and reproducibility across studies.

### Image quality has largely consistent spatial relationships with cortical morphometry

We demonstrated high correlations and similarity of spatial maps between metrics. This was particularly true for cortical thickness, which also showed the strongest associations. Notably, associations for the automatically generated Euler number were almost identical to the three manual ratings for cortical thickness, but had small but significant divergences for cortical surface area and volume. The striking spatial similarity of FSQC effects with those of motion (both here and in previous work ^2,4,5^) confirm that motion is one of the principal sources to impact image quality. With our evaluation we provide a more direct and comprehensive evaluation of image quality than motion alone, also accounting for the quality of the cortical reconstruction, another important source of bias ^14^.

Consistent with previous studies ^1,2,4,5,14,15^, we observed largely negative correlations between all three cortical phenotypes and image quality in most brain regions, with a few exceptions. In the case of motion, this is thought to be primarily due to reduced grey-white matter contrast and blurring of the cortical boundary, resulting in incorrect surface reconstruction and, typically, underestimation of cortical thickness ^2,4^. Inaccurate surface reconstruction seems to have a similar effect ^14^. Cortical volume and surface area estimates seem to be more robust to these types of errors, likely due to the fact that the GM-WM boundary is more impacted than the pial surface, and consequently SA (which relies on the GM-pial surface boundary) and volume (which is a product of SA and CT) show less of an effect of image quality ^4^. Indeed, spatial maps for cortical volume were similar to those for thickness, but with weaker relationships, and those for surface area were further attenuated still, with a few key spatial differences.

Importantly, these effects were not uniform across the cortex, with some regions being far more susceptible to image quality impacts than others, and some differing in directionality of effects, consistent with previous findings ^1,2,4,5,15^. Cortical volume and surface area largely showed similar spatial patterning, though with more positive relationships than CT, particularly for SA, and its relationship with Euler number. Some of the regions in which the strongest effects were observed, including the visual cortex, the temporal pole, and primary motor regions, are known to have unique morphometry which may render them more susceptible to issues with image quality and inaccurate surface reconstruction ^47^. The temporal and frontal poles are also regions known to have questionable signal quality ^48^. Other regional variations in the strength of relationship may in part be attributable to spatial differences in the magnitude of displacement caused by in-scanner head movement, due to participant positioning and restraints or cushioning ^5^. Another factor appears to be the thickness of the region, with higher rates of surface reconstruction errors in areas with thinner cortex causing artificially inflated thickness values ^4^. Thus, particular care should be given to interpretation of results for regions which are demonstrably susceptible to image quality.

### Thresholding analyses

Consistent with previous work ^1,2,14^, effects of quality were significantly attenuated, but not removed, when excluding participants above a certain cut-off and in a progressive thresholding manner. Excluding participants with the worst image quality may be necessary to limit the impact of bad image quality, though it may not remove its impact entirely. The progressive thresholding effects were quite similar for both FSQC and Euler. For Euler, the initial drop off in number of significant regions remaining after QC occurred more quickly but subsequently tapered off, whereas for FSQC the drop off began more gradually, but less significant regions remained after the most stringent threshold than for Euler. In the supplementary Euler percent thresholding analyses, an inflection point for the number of significant regions remaining occurs around 20%, tapering off thereafter. The decrease was more gradual with MAD thresholding. Notably, the speed of attenuation of effect size with increasing QC threshold also varied by region. Thresholding is a balancing act between decreasing the impact of noise and retaining meaningful sample representation and sufficient statistical power and thus may not be appropriate in all contexts. However, our analysis shows that even a minimal threshold can greatly improve the reliability of subsequent down-stream results.

### Image quality affects case-control differences

Importantly, the effect sizes for quality are, on average, far greater than those of diagnosis, which is concerning in light of evidence that autistic individuals (and those with other clinical diagnoses) tend to move more and have worse image quality than neurotypical controls, in our dataset as well as others ^4,5^. Thus, there is a high risk of the effects of image quality overshadowing potential diagnostic or group differences, in particular given the finding that the relationship between CT and quality was stronger in the autistic group (likely due to the greater range in quality). In our case-control comparisons, we observed subtle but significant differences depending on the extent and manner in which we controlled for image quality. Notably, when not accounting for QC in any way, some significant negative differences were observed (i.e., lower CT in autistic compared to neurotypical individuals), although not all of these survived FDR correction. After accounting for QC, these negative associations were diminished, while the positive associations (i.e., greater CT in autism than controls) were strengthened. Similar effects have been reported previously ^16^. This is unsurprising given that apparent cortical thinning is known to occur with decreased quality across much of the cortex, coupled with poorer image quality and more motion in autistic individuals. This further underscores the importance of appropriate quality control procedures for case-control analyses.

The results of the diagnosis analyses were largely consistent when controlling for FSQC or Euler at thresholds equating to approximately the same level of stringency, with only very minor differences. Results were also largely consistent when thresholding by, compared to controlling for, QC score. However, a significant difference was the emergence of significant differences (greater thickness in the autistic group than controls) in the left superior temporal gyrus when including either measure as a covariate, but not when thresholding. In the absence of a gold-standard ground truth, it is interesting to note that this is a region that has often been implicated in autism in previous work ^16,49,50^. It should also be noted that one region that is consistently significant in the case-control comparisons is the occipital cortex, which is also one of the regions in which we observe the strongest relationship with image quality. Although the effect size is attenuated once QC is accounted for, it remains significant in most of the analyses.

Little work has previously examined the impact of QC on our ability to detect group differences or alterations related to specific diagnoses or conditions. However, several reports of the impact of QC on the effects of age and trajectories of neurodevelopment ^6,14,15^ have demonstrated the potential for quality to influence relationships between neuroanatomy and demographic variables of interest. More specifically, motion and other aspects of quality have been demonstrated to both inflate and obscure relationships between age and cortical thickness, and to influence the shape of developmental trajectories ^5,6,14,15^. The effect sizes for age are typically still larger than those for quality, and therefore unlikely to completely account for previously reported age effects ^5^; however, it may lead to the exaggeration of apparent developmental effects, or ageing-related cortical thinning or atrophy. Moreover, as we have demonstrated, when it comes to diagnostic differences, effect sizes are often subtle and small compared to the relatively strong effects of motion and quality; thus, extra care and attention to QC must be paid when studying neurodevelopmental and psychiatric conditions.

### Balancing options for accounting for quality in neuroimaging studies

We have discussed and presented two main ways of accounting for quality in analyses: identifying a cut-off point and excluding all participants above or below a specific quality threshold, or controlling for quality scores by including them as a covariate in the statistical analysis. There are benefits and potential pitfalls for both options, and depending on the context one might be preferable to the other.

Excluding participants with poor image quality is a common method for QC; however, while this can ensure that the effects of quality are minimised, there are downsides to removing data. First, this necessarily results in a reduction of sample size, and consequently power, which is undesirable particularly considering the cost and effort required to collect neuroimaging data, especially in vulnerable populations. Second, and perhaps more importantly, excluding participants who are likely to have the lowest quality scans introduces unavoidable bias to the dataset: these individuals are likely to be younger and male, and to have a clinical diagnosis, more severe clinical symptoms, and lower IQ ^4,5,16^. In the context of clinical studies, this can result in samples skewed towards older participants with milder presentations and no intellectual disabilities, thereby potentially excluding participants who could benefit most from research that does not rely on verbal assessment or a minimum IQ ^51^. This bias needs to be balanced with the knowledge that poor quality data may have limited utility or lead to spurious results. Also of note is that image quality in our sample varied significantly by site, highlighting the importance of properly accounting for site effects in multi-site analyses. This also suggests that scanner hardware and sequences may contribute to quality; thus, there is unlikely to be a universal quality threshold that is applicable to all datasets, and this will need to be determined for each individual study.

An alternative solution is to retain all participants, and instead to control for QC by including quality scores as a covariate in the analysis. This avoids some of the above-mentioned biases, but introduces alternate problems. First, retaining all scans regardless of quality risks skewing results, and simply including quality as a covariate is unlikely to account for extreme values in the case of very poor quality scans. Another issue is the potential for collider bias, occurring when an independent and dependent variable both influence a third variable which is controlled for in an analysis, leading to an apparent (but spurious, or inflated) association ^52,53^. In this case, controlling for quality could influence the association between diagnosis and cortical morphometry. However, selection bias can also be considered a form of collider bias, thus this is an issue that should be taken into account regardless of the QC mitigation method chosen. Finally, to balance pros and cons and harmonise approaches, a hybrid solution can be implemented, whereby only the worst scans which are considered unusable are excluded, and QC is included in the model to correct for any residual effects caused by other lower quality, but still potentially usable, scans.

### Limitations

These results should be interpreted in light of certain limitations. First, no quality metric is perfect, and as mentioned above there is no gold standard. Without prospective motion trackers installed at the time of scanning, we cannot accurately quantify motion, and all visual inspections of scan and surface reconstruction quality will have some level of subjectivity. We attempt to mitigate this by comparing multiple QC metrics, both automated and manually rated, by multiple independent raters. Next, we rely on two metrics, FSQC and Euler number, which are specific to FreeSurfer, and thus may have limited generalisability. However, our FSQC tool could easily be applied to other processing and surface reconstruction tools. Other automated metrics of image quality exist (e.g., MRI-QC, Qoala-T), but were not evaluated in the current study. We also focused exclusively on cortical morphometry. Given recent evidence that subcortical structures are also influenced by quality (though potentially to a lesser degree) ^1^, extending the current work to subcortical structures, particularly in the context of clinical group differences, could be valuable. Finally, our sample, the ABIDE dataset, consists of a relatively limited demographic, including mostly children and young adults, a substantial proportion of whom have a diagnosis of autism. However, this dataset allowed us to examine the impact of quality on case-control differences, and we successfully replicated at least some of our results in a much larger, more representative sample.

### Conclusion

Our results highlight the importance of careful quality control of neuroimaging data, and some of the potential consequences of failing to do so. We explored the effect of various QC metrics and mitigation techniques, and demonstrated that these can have a significant impact on our ability to detect differences in neuroanatomy related to autism.

## Data Availability

The ABIDE dataset is available to download at: https://fcon_1000.projects.nitrc.org/indi/abide/
The imaging rating tool, code to generate QC png images and analysis scripts are available at:
https://github.com/sbedford0/FSQC.
The full protocol can be found at: https://dx.doi.org/10.17504/protocols.io.kxygx9m6wg8j/v1

## Acknowledgements

Data were curated and analysed using a computational facility funded by an MRC research infrastructure award (MR/M009041/1) to the School of Clinical Medicine, University of Cambridge and supported by the mental health theme of the NIHR Cambridge Biomedical Research Centre. The views expressed are those of the authors and not necessarily those of the NIH, NHS, the NIHR or the Department of Health and Social Care.

## References

1. Gilmore, A. D., Buser, N. J. & Hanson, J. L. Variations in structural MRI quality significantly impact commonly used measures of brain anatomy. Brain informatics 8, (2021).

2. Reuter, M. et al. Head motion during MRI acquisition reduces gray matter volume and thickness estimates. Neuroimage 107, 107–115 (2015).

3. Tisdall, M. D. et al. Prospective motion correction with volumetric navigators (vNavs) reduces the bias and variance in brain morphometry induced by subject motion. Neuroimage 127, 11–22 (2016).

4. Pardoe, H. R., Kucharsky Hiess, R. & Kuzniecky, R. Motion and morphometry in clinical and nonclinical populations. Neuroimage 135, 177–185 (2016).

5. Alexander-Bloch, A. et al. Subtle in-scanner motion biases automated measurement of brain anatomy from in vivo MRI. Hum. Brain Mapp. 2397, 2385–2397 (2016).

6. Savalia, N. K. et al. Motion-related artifacts in structural brain images revealed with independent estimates of in-scanner head motion. Hum. Brain Mapp. 38, 472–492 (2017).

7. Makowski, C., Lepage, M. & Evans, A. C. Head motion: the dirty little secret of neuroimaging in psychiatry. J. Psychiatry Neurosci. 44, 62–68 (2019).

8. Satterthwaite, T. D. et al. Impact of in-scanner head motion on multiple measures of functional connectivity: Relevance for studies of neurodevelopment in youth. Neuroimage 60, 623–632 (2012).

9. van Dijk, K. R. A., Sabuncu, M. R. & Buckner, R. L. The influence of head motion on intrinsic functional connectivity MRI. Neuroimage 59, 431–438 (2012).

10. Power, J. D., Barnes, K. A., Snyder, A. Z., Schlaggar, B. L. & Petersen, S. E. Spurious but systematic correlations in functional connectivity MRI networks arise from subject motion. Neuroimage 59, 2142–2154 (2012).

11. Goto, M. et al. Head Motion and Correction Methods in Resting-state Functional MRI. Magn. Reson. Med. Sci. 15, 178–186 (2016).

12. Baum, G. L. et al. The impact of in-scanner head motion on structural connectivity derived from diffusion MRI. Neuroimage 173, 275–286 (2018).

13. Bastiani, M. et al. Automated quality control for within and between studies diffusion MRI data using a non-parametric framework for movement and distortion correction. Neuroimage 184, 801–812 (2019).

14. Ducharme, S. et al. Trajectories of cortical thickness maturation in normal brain development--The importance of quality control procedures. Neuroimage 125, 267–279 (2016).

15. Rosen, A. F. G. et al. Quantitative assessment of structural image quality. Neuroimage 169, 407–418 (2018).

16. Bedford, S. A. et al. Large-scale analyses of the relationship between sex, age and intelligence quotient heterogeneity and cortical morphometry in autism spectrum disorder. Mol. Psychiatry 25, 614–628 (2020).

17. Yendiki, A., Koldewyn, K., Kakunoori, S., Kanwisher, N. & Fischl, B. Spurious group differences due to head motion in a diffusion MRI study. Neuroimage 88, 79–90 (2014).

18. Bethlehem, R. A. I. et al. Brain charts for the human lifespan. Nature 604, 525–533 (2022).

19. David C. Van Essen, M. F. G. The Human Connectome Project: Progress and Prospects. Cerebrum 2016, (2016).

20. Postema, M. C. et al. Altered structural brain asymmetry in autism spectrum disorder in a study of 54 datasets. Nat. Commun. 10, 1–12 (2019).

21. Di Martino, A. et al. Enhancing studies of the connectome in autism using the autism brain imaging data exchange II. Sci Data 4, 170010 (2017).

22. Weiner, M. W. et al. Impact of the Alzheimer’s Disease Neuroimaging Initiative, 2004 to 2014. Alzheimers. Dement. 11, 865–884 (2015).

23. Thompson, P. M. et al. The ENIGMA Consortium: large-scale collaborative analyses of neuroimaging and genetic data. Brain Imaging Behav. 8, 153–182 (2014).

24. Marek, S. et al. Reproducible brain-wide association studies require thousands of individuals. Nature 603, 654–660 (2022).

25. Ai, L. et al. Is it time to switch your T1W sequence? Assessing the impact of prospective motion correction on the reliability and quality of structural imaging. Neuroimage 226, 117585 (2021).

26. Esteban, O. et al. MRIQC: Advancing the automatic prediction of image quality in MRI from unseen sites. PLoS One 12, (2017).

27. Klapwijk, E. T., van de Kamp, F., van der Meulen, M., Peters, S. & Wierenga, L. M. Qoala-T: A supervised-learning tool for quality control of FreeSurfer segmented MRI data. Neuroimage 189, 116–129 (2019).

28. Keshavan, A., Yeatman, J. D. & Rokem, A. Combining Citizen Science and Deep Learning to Amplify Expertise in Neuroimaging. Front. Neuroinform. 13, 29 (2019).

29. Backhausen, L. L. et al. Quality Control of Structural MRI Images Applied Using FreeSurfer-A Hands-On Workflow to Rate Motion Artifacts. Front. Neurosci. 10, (2016).

30. Raamana, P. R. et al. Visual QC Protocol for FreeSurfer Cortical Parcellations from Anatomical MRI. bioRxiv 10, 2020.09.07.286807 (2021).

31. Protocol for Quality Control and Summary Statistics " ENIGMA. https://enigma.ini.usc.edu/protocols/imaging-protocols/protocol-for-quality-control-and-summary-statistics/.

32. Di Martino, A. et al. The autism brain imaging data exchange: towards a large-scale evaluation of the intrinsic brain architecture in autism. Mol. Psychiatry 19, 659–667 (2014).

33. Haar, S., Berman, S., Behrmann, M. & Dinstein, I. Anatomical Abnormalities in Autism? Cereb. Cortex 26, 1440–1452 (2016).

34. Schaer, M., Kochalka, J., Padmanabhan, A., Supekar, K. & Menon, V. Sex differences in cortical volume and gyrification in autism. Mol. Autism 6, 42 (2015).

35. Valk, S. L., Di Martino, A., Milham, M. P. & Bernhardt, B. C. Multicenter mapping of structural network alterations in autism. Hum. Brain Mapp. 36, 2364–2373 (2015).

36. Bethlehem, R. A. I., Romero-Garcia, R., Mak, E., Bullmore, E. T. & Baron-Cohen, S. Structural covariance networks in children with autism or ADHD. Cereb. Cortex 27, 4267– 4276 (2017).

37. Bethlehem, R. A. I. et al. A normative modelling approach reveals age-atypical cortical thickness in a subgroup of males with autism spectrum disorder. Commun Biol 3, 486 (2020).

38. Khundrakpam, B. S., Lewis, J. D., Kostopoulos, P., Carbonell, F. & Evans, A. C. Cortical Thickness Abnormalities in Autism Spectrum Disorders Through Late Childhood, Adolescence, and Adulthood: A Large-Scale MRI Study. Cereb. Cortex 27, 1721–1731 (2017).

39. Kohli, J. S. et al. Local Cortical Gyrification is Increased in Children With Autism Spectrum Disorders, but Decreases Rapidly in Adolescents. Cereb. Cortex 29, 2412–2423 (2019).

40. Turner, A. H., Greenspan, K. S. & van Erp T. G. M. Pallidum and lateral ventricle volume enlargement in autism spectrum disorder. Psychiatry Res Neuroimaging 252, 40–45 (2016).

41. Floris, D. L., Lai, M.-C., Nath, T., Milham, M. P. & Di Martino, A. Network-specific sex differentiation of intrinsic brain function in males with autism. Mol. Autism 9, 17 (2018).

42. Olafson, E. et al. Examining the Boundary Sharpness Coefficient as an Index of Cortical Microstructure in Autism Spectrum Disorder. Cereb. Cortex 31, 3338–3352 (2021).

43. Nielsen, J. A. et al. Abnormal lateralization of functional connectivity between language and default mode regions in autism. Mol. Autism 5, 8 (2014).

44. Ray, S. et al. Structural and functional connectivity of the human brain in autism spectrum disorders and attention-deficit/hyperactivity disorder: A rich club-organization study. Hum. Brain Mapp. 35, 6032–6048 (2014).

45. Glasser, M. F. et al. A multi-modal parcellation of human cerebral cortex Europe PMC Funders Group. Nature 536, 171–178 (2016).

46. Desikan, R. S. et al. An automated labeling system for subdividing the human cerebral cortex on MRI scans into gyral based regions of interest. (2006) doi:10.1016/j.neuroimage.2006.01.021.

47. Scholtens, L. H., de Reus, M. A. & van den Heuvel, M. P. Linking contemporary high resolution magnetic resonance imaging to the von Economo legacy: A study on the comparison of MRI cortical thickness and histological measurements of cortical structure. Hum. Brain Mapp. 36, 3038–3046 (2015).

48. McCarthy, C. S. et al. A comparison of FreeSurfer-generated data with and without manual intervention. Front. Neurosci. 9, 379 (2015).

49. Jou, R. J., Minshew, N. J., Keshavan, M. S., Vitale, M. P. & Hardan, A. Y. Enlarged right superior temporal gyrus in children and adolescents with autism. Brain Res. 1360, 205–212 (2010).

50. Ecker, C. The neuroanatomy of autism spectrum disorder: An overview of structural neuroimaging findings and their translatability to the clinical setting. Autism 21, 18–28 (2017).

51. Nordahl, C. W. et al. Methods for acquiring MRI data in children with autism spectrum disorder and intellectual impairment without the use of sedation. J. Neurodev. Disord. 8, 20 (2016).

52. Holmberg, M. J. & Andersen, L. W. Collider Bias. JAMA 327, 1282–1283 (2022).

53. Munafò, M. R., Tilling, K., Taylor, A. E., Evans, D. M. & Davey Smith, G. Collider scope: when selection bias can substantially influence observed associations. Int. J. Epidemiol. 47, 226–235 (2018).

