## Supplementary methods and materials for "The impact of quality control on cortical morphometry comparisons in autism"

**Table of contents**

**Supplementary methods**	**1**

1. Inter-rater reliability	1

1.1 Comparison of rater maps	1

2. Additional main analyses	1

2.1 Desikan-Killiany atlas results	1

2.2 Meta-analysis results	1

2.3 Replication in Lifespan sample	1

2.4 Variance partitioning	2

3. Thresholding analyses	2

3.1 Euler thresholding per site	2

3.2 FSQC median split analysis	2

3.3 Percent exclusion analysis	2

4. Diagnosis	3

4.1 Cut off points	3

4.2 Thresholding by FSQC but controlling for Euler	3

4.3 Diagnosis Interaction	3

4.4 SA and CV diagnosis analysis	3

**Supplementary results**	**4**

1. Inter-rater reliability	4

1.1 SB, RB and average scores	4

2. Sensitivity analyses (main results)	5

2.1 DK results	6

2.2 Meta-analysis results	7

2.3 Replication in Lifespan sample	9

2.4 Variance partitioning	9

3. Thresholding analyses	13

3.1 Euler threshold per site (CT)	13

3.2 FSQC median split analysis (CT)	14

3.3 Percent exclusion analysis	15

3.4 SA and CV: FSQC thresholding	19

3.5 SA and CV: Euler thresholding	20

4. Diagnosis	22

4.1 Diagnosis with cut off points	22

4.2 Diagnosis FSQC threshold + Euler	23

Fig S4.2. Effect of diagnosis on cortical thickness at various FSQC thresholds and controlling for Euler number.	23

4.3 Diagnosis interaction	23

4.4 Diagnosis SA and CV	24

### Supplementary methods

#### 1. Inter-rater reliability

##### 1.1 Comparison of rater maps

To ensure results did not differ by rater, the same linear mixed effects models were conducted on cortical thickness using FSQC scores by each main rater (SB, RB), as well as the average score between both raters, and maps displaying the results across the cortex were visually compared across the three analyses.

#### 2. Additional main analyses

##### 2.1 Desikan-Killiany atlas results

For comparison with previous work and our replication analysis, linear mixed effects models were run for each Desikan-Killiany parcellation, using the same models as the main analyses, for CT, SA and CV, and partial correlations extracted and plotted. Results were corrected for multiple comparisons using the false discovery rate (FDR) across parcellations in all analyses.

##### 2.2 Meta-analysis results

The FSQC and Euler analyses for cortical thickness were also conducted using a meta-analytic approach, as an alternative method to mixed effects models for controlling for site differences. This method also provides information about intersite heterogeneity. For these analyses, linear models were run separately per site, including QC metric, age, age^2^, and sex, and then pooled across sites in a random effects meta-analysis based on partial r correlation coefficients, using the metafor package in R.

##### 2.3 Replication in Lifespan sample

To validate and attempt to replicate our results in a larger, more representative dataset covering the whole human lifespan (0-100 years), we ran the same main analyses for Euler using one of the largest MRI datasets currently available (see Bethlehem et al, 2022). To avoid confounds of various diagnoses, we used controls only from this dataset, and excluded ABIDE so as not to have overlap with our original analysis, resulting in a sample of 74,647 individuals. We then ran the same linear mixed effects models, including Euler, age, age^2^ and sex, with site as a random variable, for all three cortical phenotypes separately. This was done for Euler number, and using DK parcellations, as this was the data available for this dataset.

##### 2.4 Variance partitioning

We also conducted a variance partitioning analysis to quantify the proportion of variance accounted for by scan quality (FSQC and Euler), relative to other covariates on global brain measures (cortical GMV, WMV, subcortical GMV, ventricular volume, total GMV, estimated TIV, and mean cortical thickness), and for cortical thickness. This analysis was conducted using the variancePartition package in R (<http://bioconductor.org/packages/variancePartition>; (Hoffman & Schadt, 2016), often used for, and adapted from, gene expression analysis. Linear mixed models are used to quantify variance in (here) cortical thickness parcellations contributed by FSQC, Euler, and other main variables: diagnosis, sex, age, and site. Percent of variance explained is then plotted for each neuroanatomical phenotype of interest (cortical and subcortical total grey matter volume, white matter volume, and ventricle volume).

#### 3. Thresholding analyses

##### 3.1 Euler thresholding per site

Due to the significant variation (in scanner, scanning parameters, demographics, and quality) between sites in the ABIDE dataset, we repeated the Euler threshold analyses applying the cut-off point based on MADs to each site individually, rather than the whole sample, for cortical thickness. MAD was calculated individually for each site, and used to exclude participants above that value for each site, at MADs of 1, 2 and 3. The same linear models as above were then run at each cut off point, assessing the relationship between CT and Euler.

##### 3.2 FSQC median split analysis

We also compared CT between participants with the best and worst scan quality, based on FSQC scores: participants were split into two groups (“high” and “low” scores) based on a median split of 1.1. A mixed linear effects model was then run, including group (high/low), age, age^2^, and sex as fixed effects, and site as a random effect.

##### 3.3 Percent exclusion analysis

Next, we wanted to assess the impact of excluding participants with the worst scan quality, based on the top percentage rather than predefined cut off points. We conducted this analysis based on Euler number, for CT, at thresholds in increments of 5%, from 5-50%, both of the whole dataset, and per individual site. As in previous analyses, regional relationships for CT were assessed at each threshold by running the same linear mixed effects models. The relationship between the number of significant regions remaining and the percentage of participants excluded was plotted, as was the relationship between the strength of the partial correlation of the strongest regions and the percent of excluded participants, to examine how excluding different proportions of the dataset impacts or attenuates the impact of scan quality on morphometric estimates.

#### 4. Diagnosis

##### 4.1 Cut off points

To further explore the impact of thresholding by scan quality, we reran the same analysis exploring the impact of diagnosis on CT at cut-off points of 3, 2.5, 2, 1.5 for FSQC, and 1, 2 and 3 MADs for Euler number. At each threshold, the model included diagnosis, age, age^2^, and sex as fixed variables and site as a random variable.

##### 4.2 Thresholding by FSQC but controlling for Euler

Finally, we examined the effect of combining FSQC and Euler as QC methods, by examining the effect of diagnosis while controlling for Euler at various FSQC thresholds (3, 2.5, 2, and 1.5 as above). Here, the model included diagnosis, age, age^2^, Euler number and sex as fixed variables and site as a random variable.

##### 4.3 Diagnosis Interaction

Finally, the interaction between diagnosis and FSQC or Euler was assessed on each cortical phenotype. For significant regions, the effect of FSQC and Euler on cortical estimates were then examined separately in the autistic and control groups.

##### 4.4 SA and CV diagnosis analysis

The main diagnosis analyses were also run for SA and CV, including examining the impact of diagnosis alone, diagnosis controlling for FSQC and Euler, and the impact of diagnosis while thresholding by each FSQC (at 2.5) and Euler (at 2 MADs).

### Supplementary results

#### 1. Inter-rater reliability

##### 1.1 SB, RB and average scores

Regional results depicting the relationship between FSQC and cortical thickness were almost identical for both raters and average ratings across the cortex.

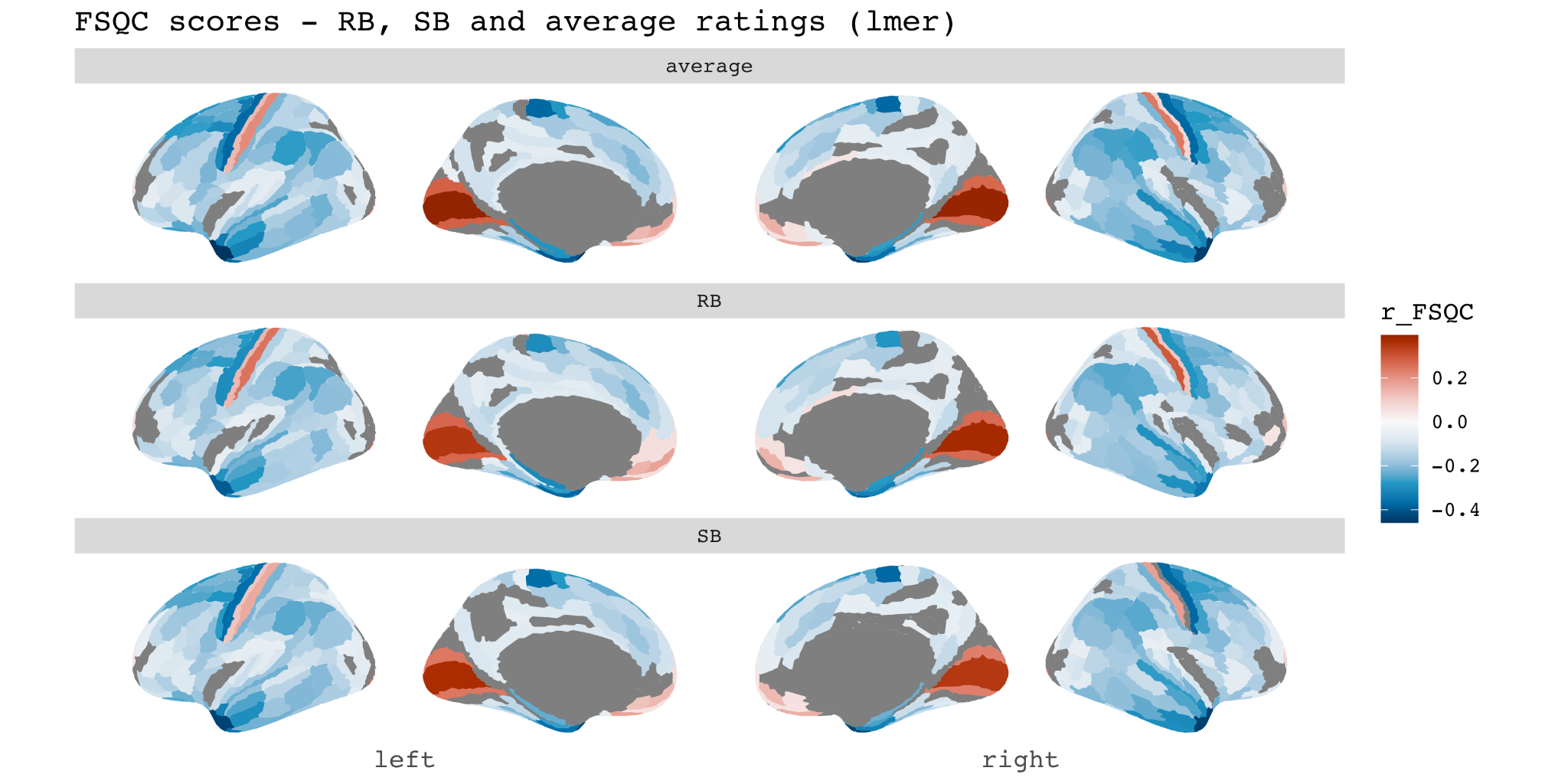

Figure S1.1 Associations between FSQC and cortical thickness for average scores between SB and RB (top), RB only (middle) and SB only (bottom row). Spatial maps were consistent across all three sets of scores.

#### 2. Sensitivity analyses (main results)

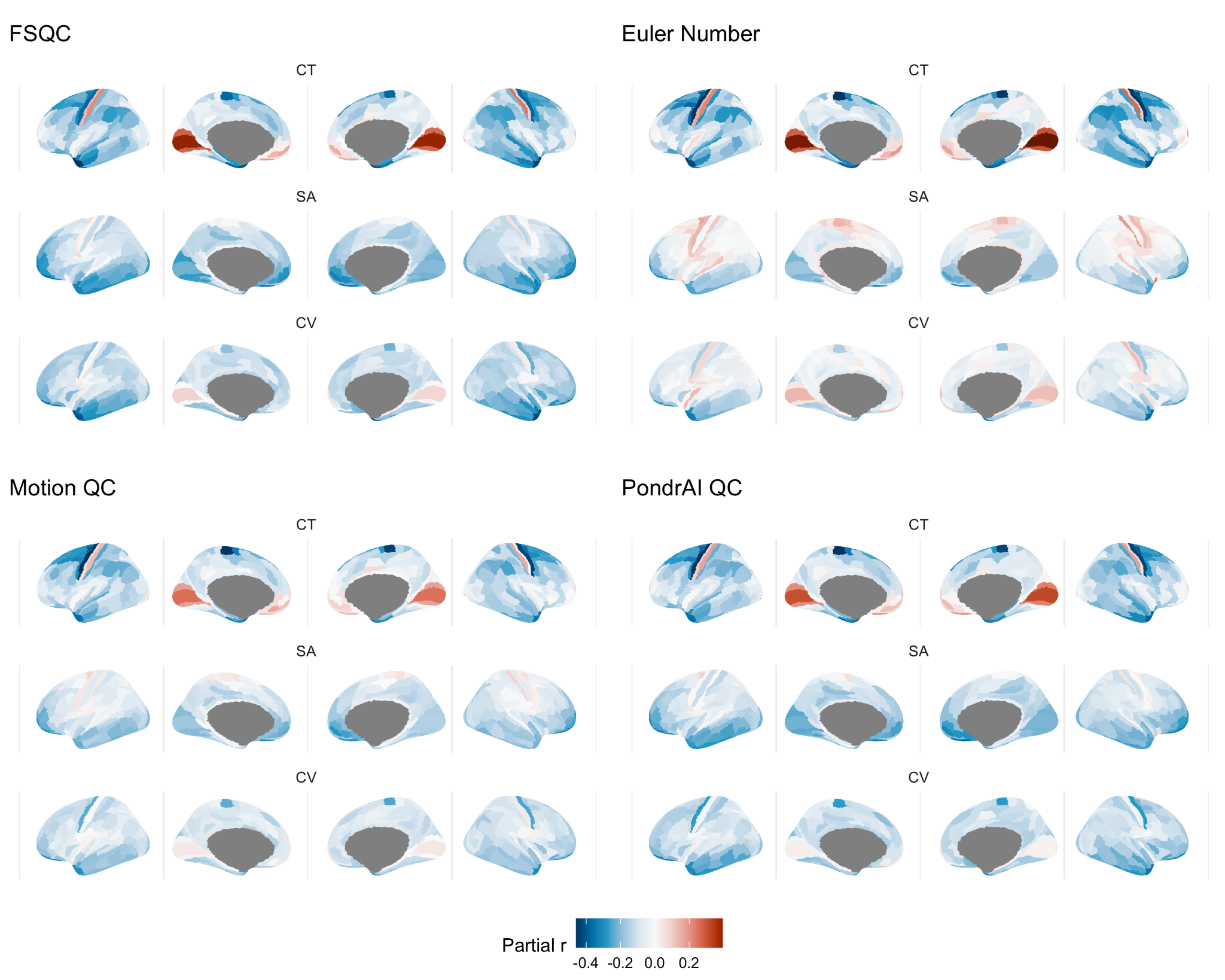

Figure S2. Results of regional analyses (main figure 2): effect of all QC metrics on each cortical phenotype, here unthresholded for significance.

**FSQC correlations with global brain measures**

|  | Spearman rho | p-value |
| --- | --- | --- |
| cGMV | -0.066 | 0.004 |
| WMV | -0.21 | < 0.0001 |
| sGMV | -0.065 | 0.005 |
| Ventricles | -0.055 | 0.018 |
| TBV | -0.157 | < 0.0001 |
| meanCT | -0.044 | 0.059 |

Supplementary table S1

##### 2.1 DK results

Results using the Desikan-Killiany parcellations were consistent with the main analyses using HCP parcellations, and again consistent across QC metrics with the exception of Euler for SA and CV. Strongest effects were seen for cortical thickness across all metrics. Associations were largely negative, with positive correlations observed in the occipital and ventromedial prefrontal cortices for CT, and motor cortex for SA.

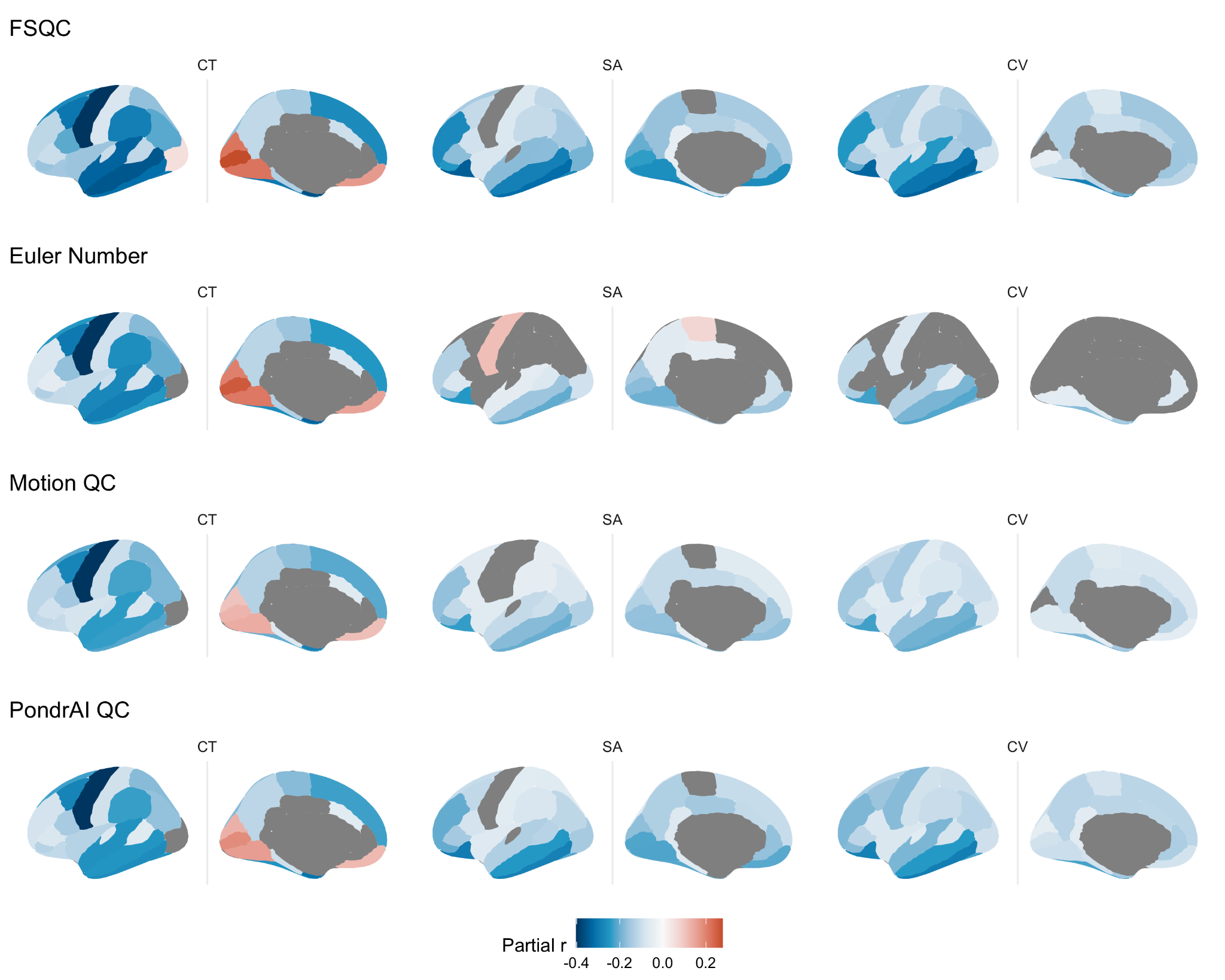

Figure S2.1. Associations between QC metrics and regional cortical morphometry using the Desikan-Killiany atlas parcellations. Results are consistent with those derived using the HCP-Glasser parcellation.

##### 2.2 Meta-analysis results

Results from the meta-analytic technique for CT also yielded consistent, though slightly weaker, results to those using linear mixed effects models. Forest plots demonstrate largely consistent results across sites, with some variability.

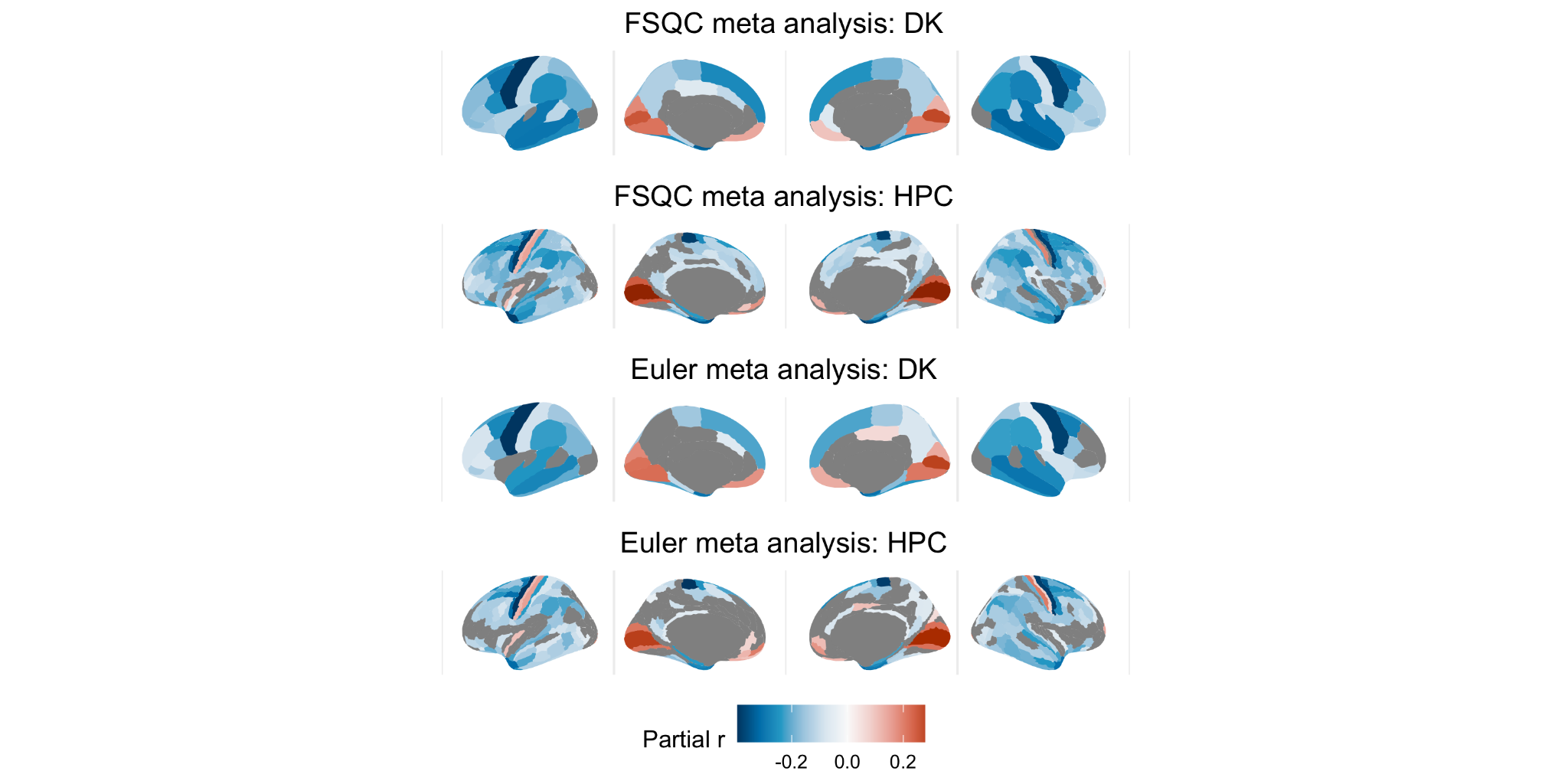

Fig. S2.2.1 Associations between Euler/FSQC and CT using a meta-analytic technique, with DK and HPC parcellations. Results are largely consistent with results from linear mixed effects models.

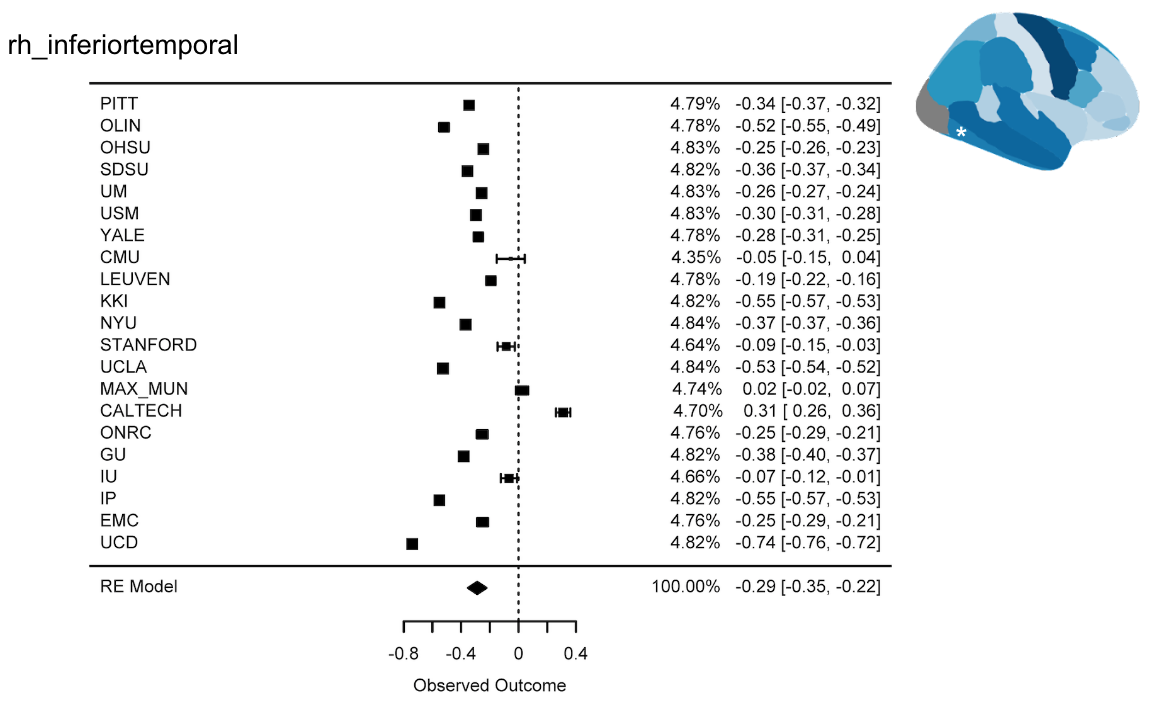

Fig S2.2.2. FSQC: Forest plot for DK inferior temporal gyrus parcellation. Effects are largely consistent across sites: all but two sites show a negative effect size, with some variability.

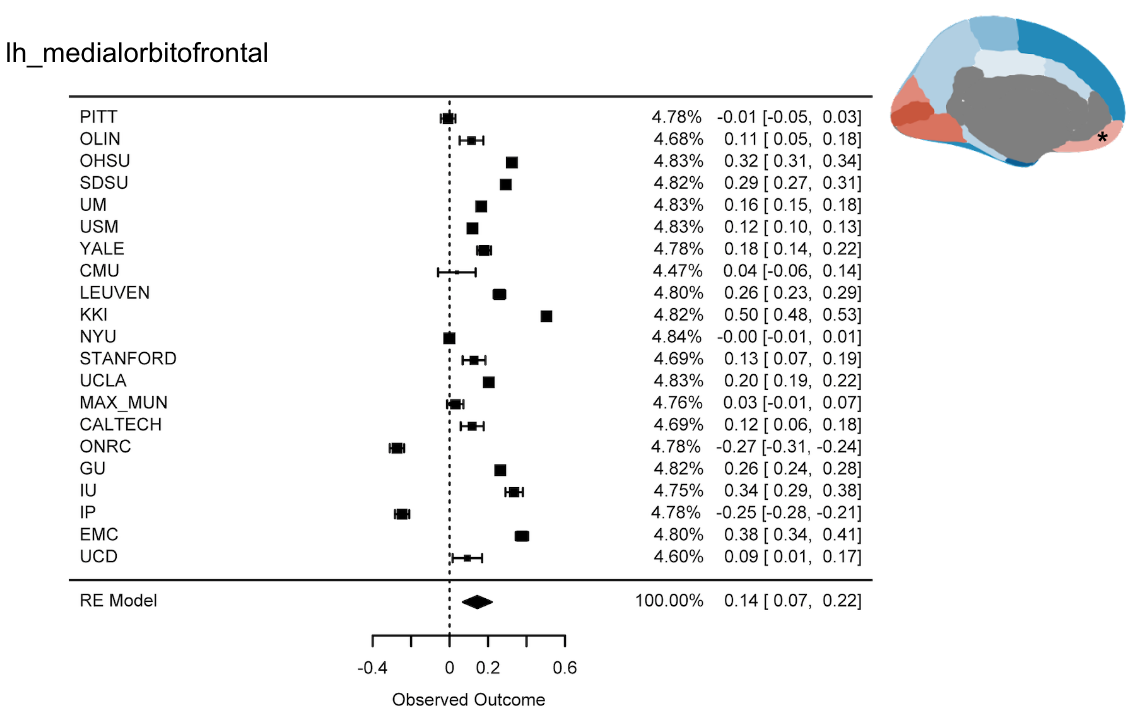

Fig S2.2.3. FSQC: Forest plot for DK ventromedial prefrontal cortex parcellation. Most sites show a positive effect size, with a few exceptions and slightly more variability.

##### 2.3 Replication in Lifespan sample

Conducting the same analyses for Euler on all three cortical phenotypes in a much larger, more representative sample yielded largely similar results with some key differences. Results for CT were almost identical, with negative associations across the cortex with the exception of the occipital and ventromedial prefrontal cortices. Results for SA differed the most, with positive associations observed across much of the cortex in this larger sample, but only in the motor cortex in the ABIDE sample. CV results were similar in magnitude and direction, but more regions reached significance in the larger sample than in ABIDE.

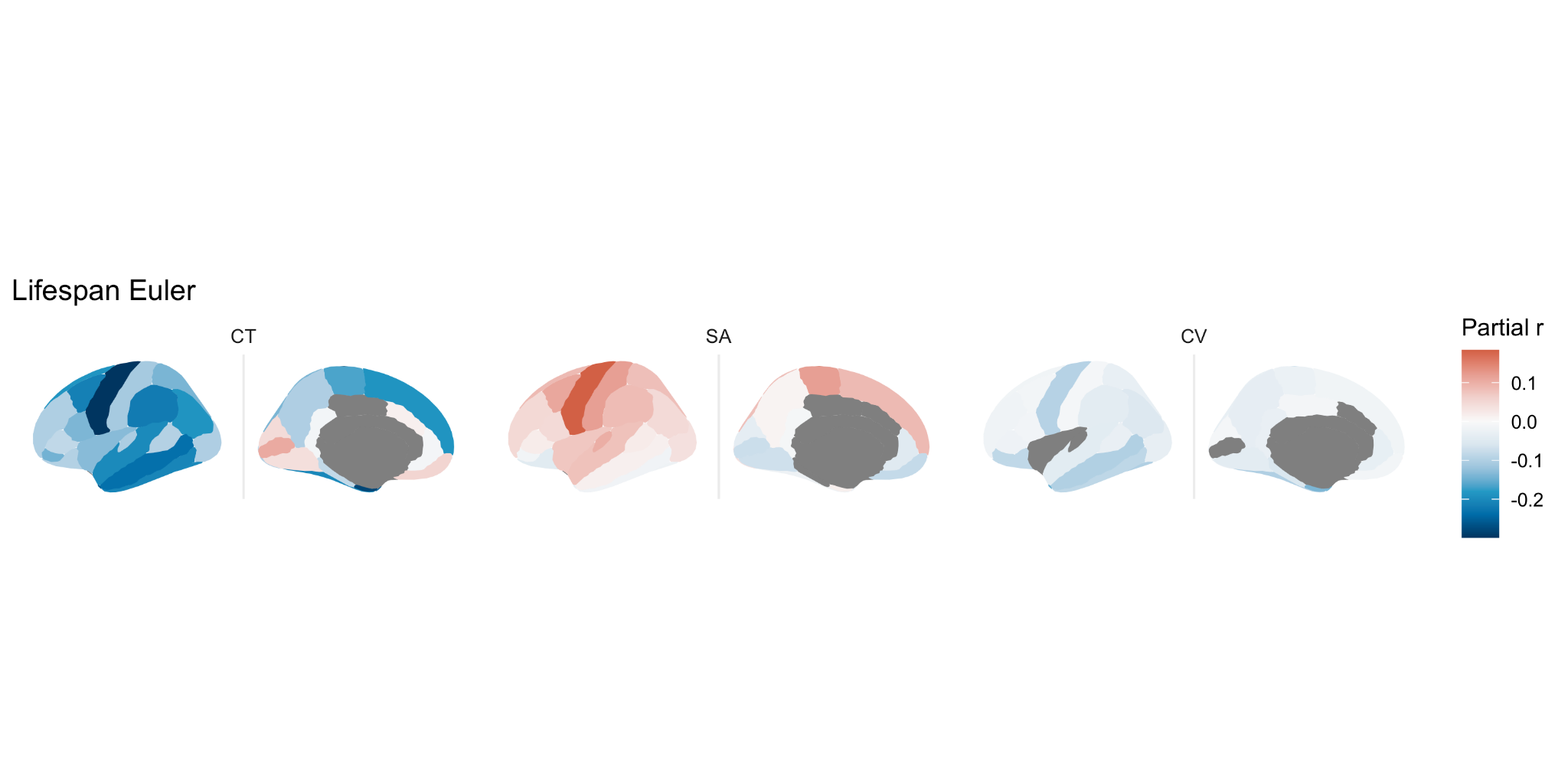

Fig S2.3. Relationship between each cortical phenotype and Euler number across the cortex in a large lifespan dataset. Results are largely consistent with ABIDE results, with the exception of more regions showing a positive relationship for SA.

##### 2.4 Variance partitioning

The variance partitioning analysis on global brain measures indicated that, on average across all global brain measures, FSQC explained 3.7%, Euler number about 1%, and diagnosis 0.2% of the variance (Figure S2.4.1). For most measures, FSQC and Euler explained only a very small amount of the variance (<2%), but more substantial proportions of FSQC for cGMV (8.7%), tGMV (7.8%), and WMV (5.3%) (Figure S2.4.2). For all measures except ventricular volume, both quality metrics contributed to a substantially larger portion of the variance than diagnosis.

Across cortical thickness HPC parcellations, FSQC explained on average 1.4% of the variance, Euler 0.8% and diagnosis less than 0.1%. Site (16%) and age (11%) explained the most variance (Figure S2.4.3). A sample of 50 parcellations and their relative contributions is shown in Figure S2.4.4; FSQC and Euler appeared to explain significantly more variance than diagnosis for almost every parcellation. Brain maps of the percent of variance explained by FSQC and Euler are shown in Figure S2.4.5.

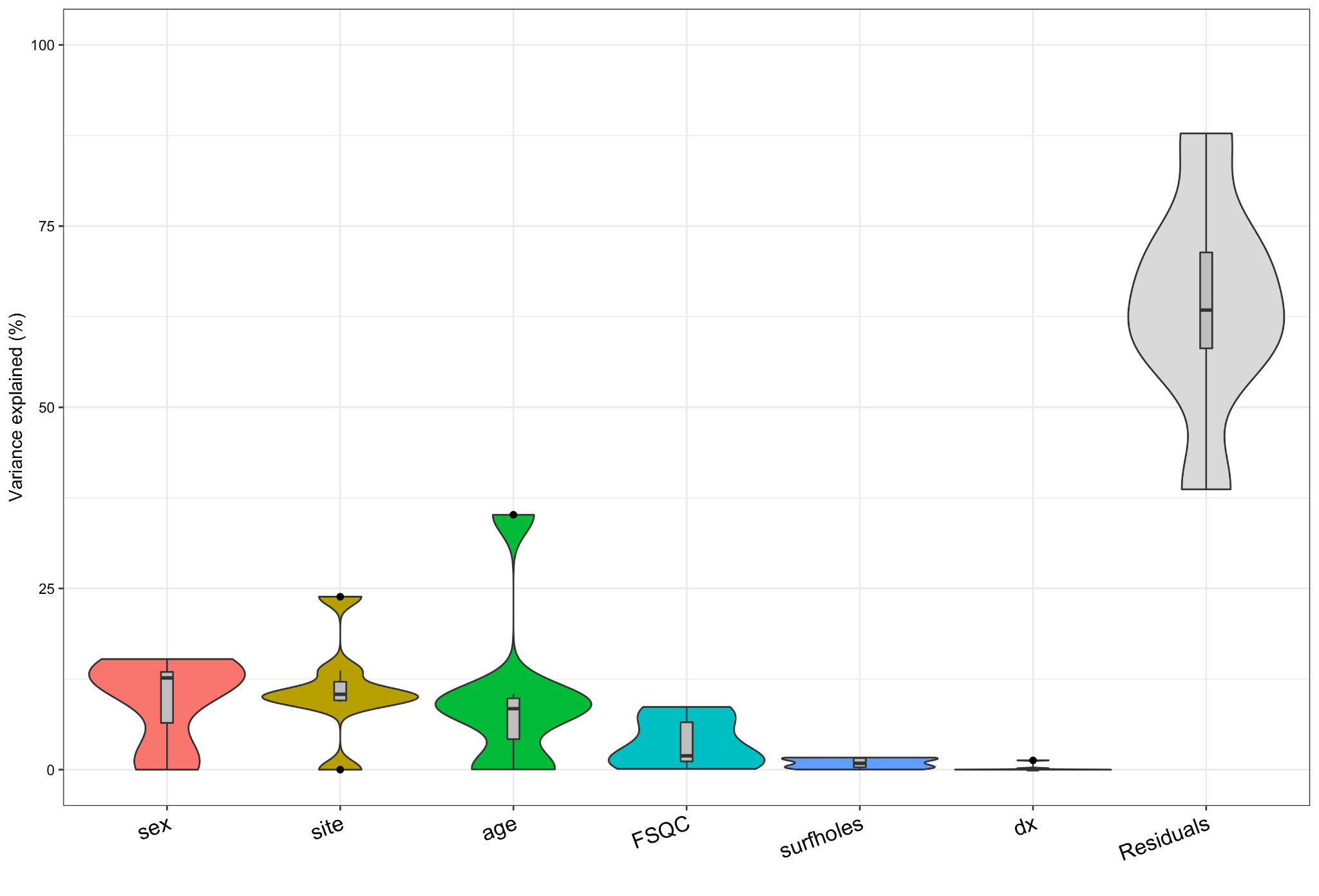

Fig S2.4.1. Average variance explained by each variable, across all global brain measures.

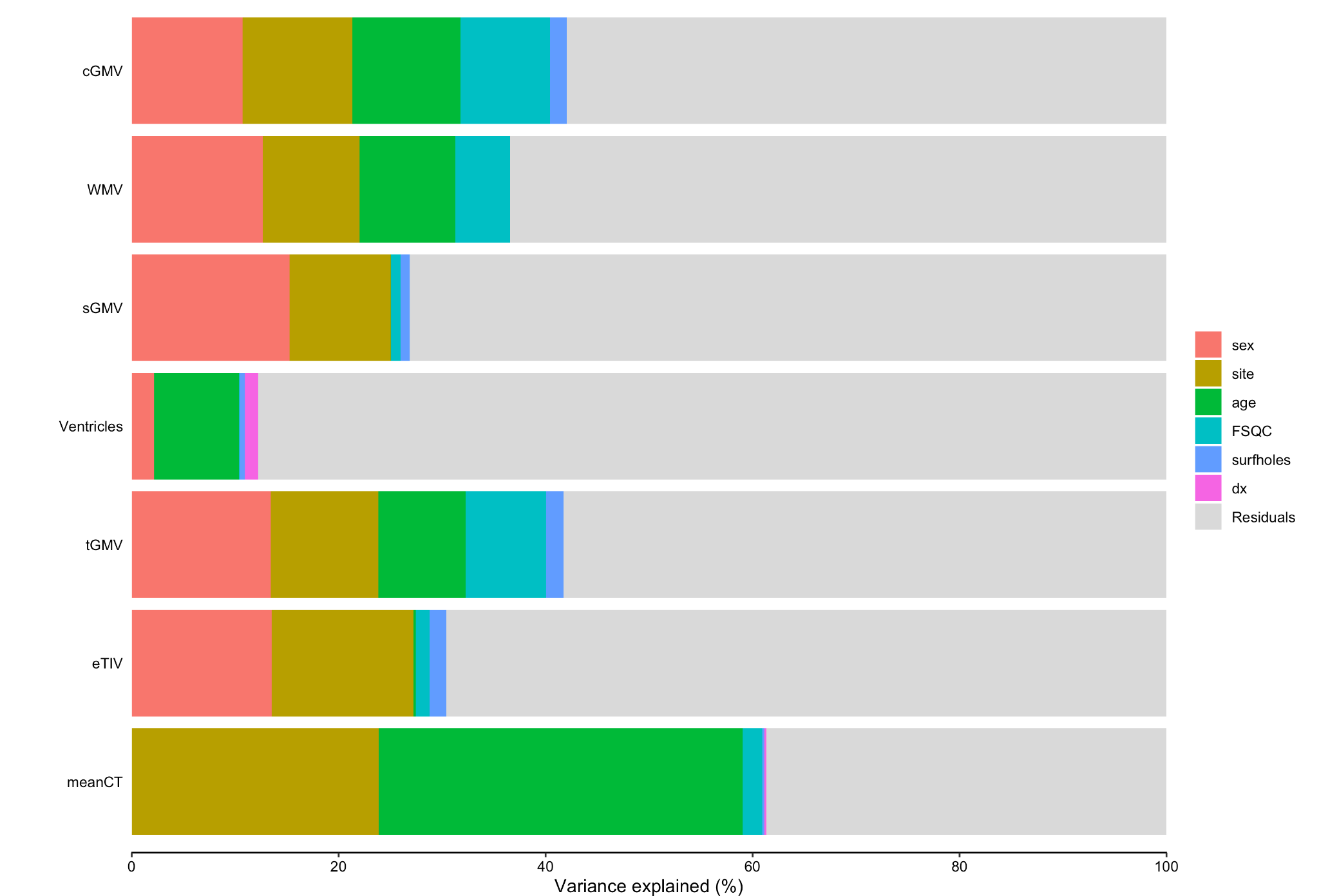

Fig S2.4.2. Percent of variance explained by each variable for each global brain measure.

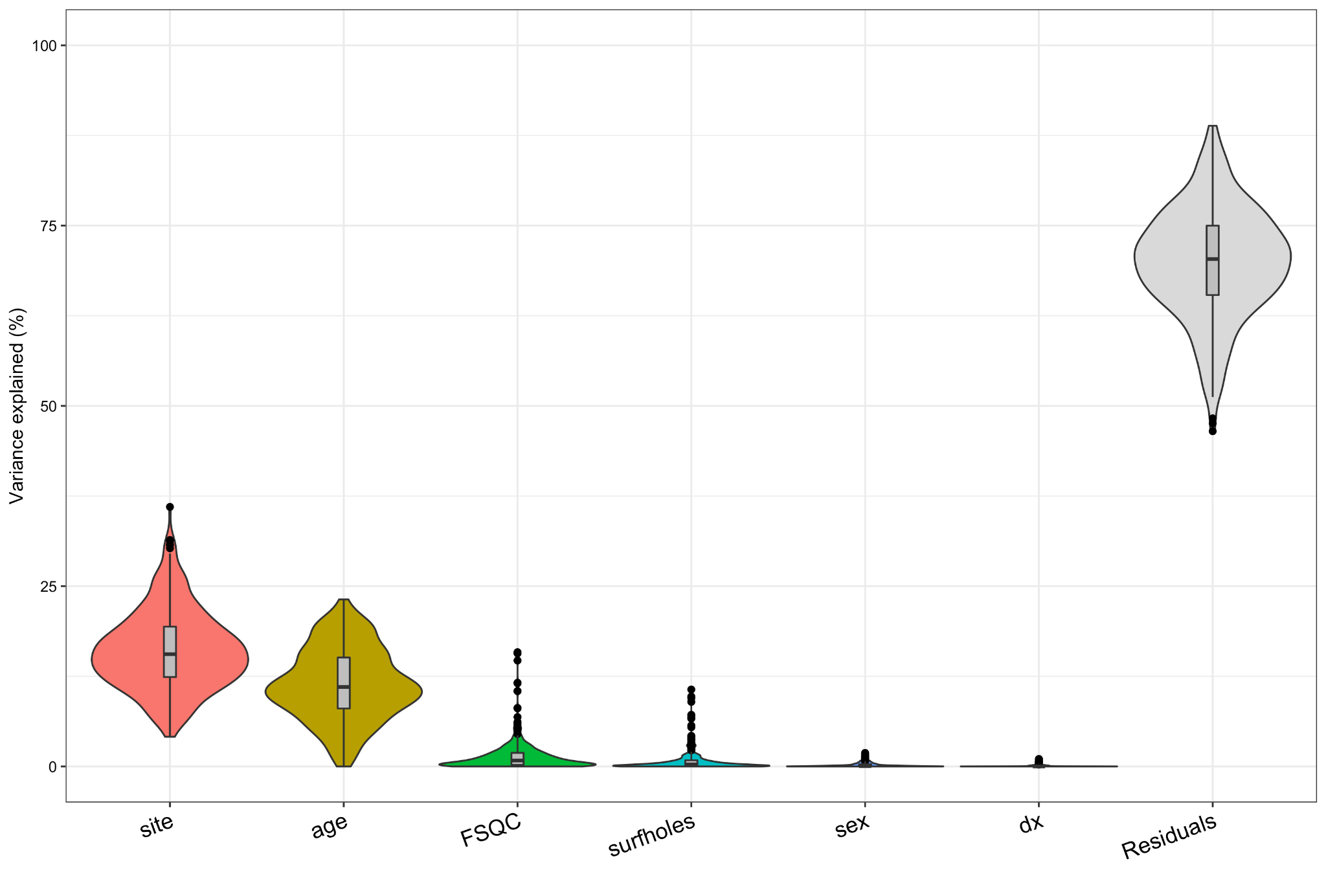

Fig S2.4.3 average variance explained by each variable, across all cortical thickness parcellations.

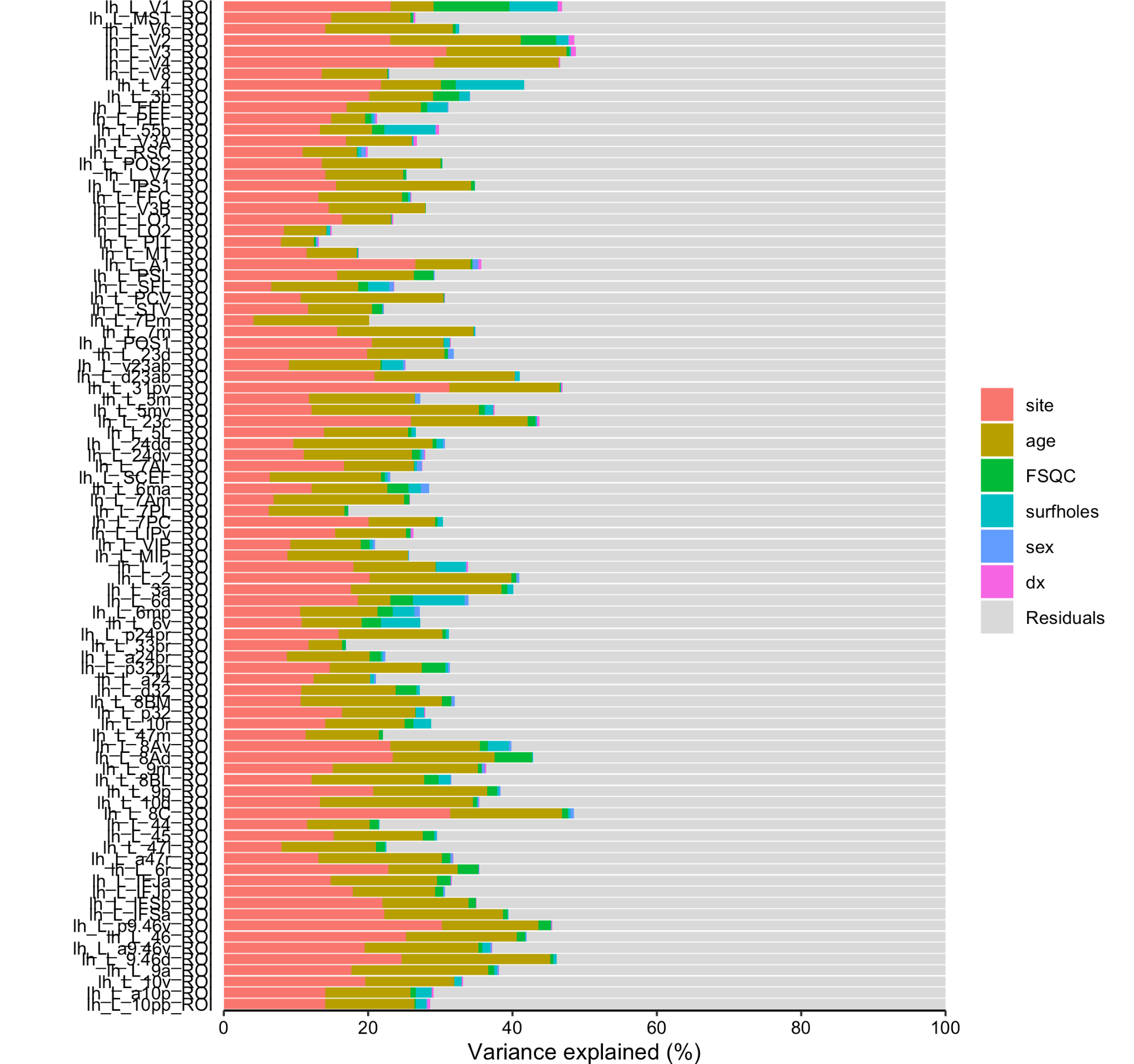

Fig S2.4.4 Percent of variance explained by each variable for the first 50 listed HPC parcellations.

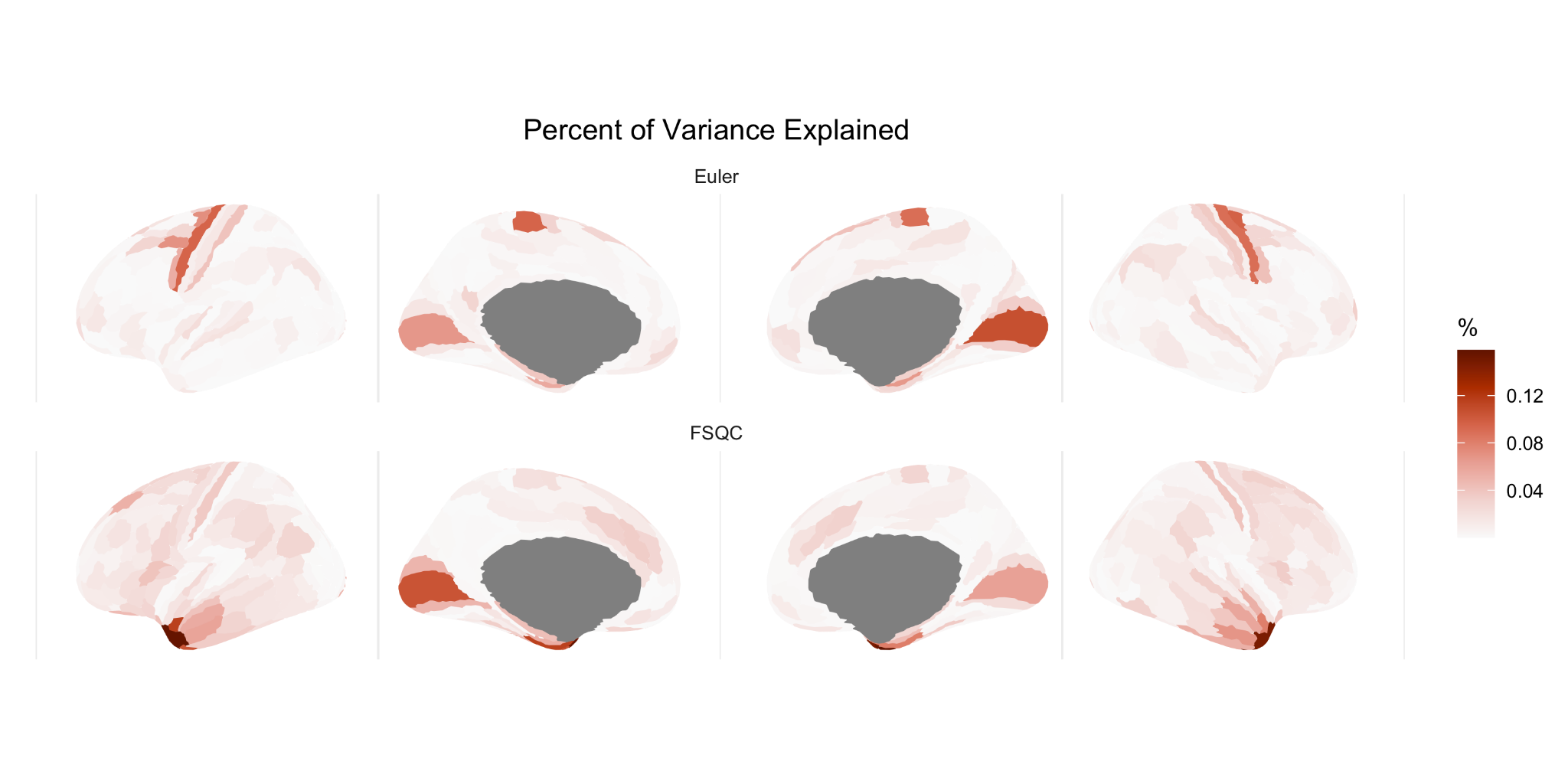

Fig S2.4.5. Percent of variance explained by Euler (top) and FSQC (bottom) for each HPC cortical thickness parcellation across the cortex.

#### 3. Thresholding analyses

##### 3.1 Euler threshold per site (CT)

When calculating and applying the MAD-based Euler threshold to each site individually rather than to the whole sample, results were comparable but with a few notable differences. Generally speaking, less regions remained significant at each cut-off point when applying the MAD-based threshold to each site individually compared to the whole sample; however, differences were minimal.

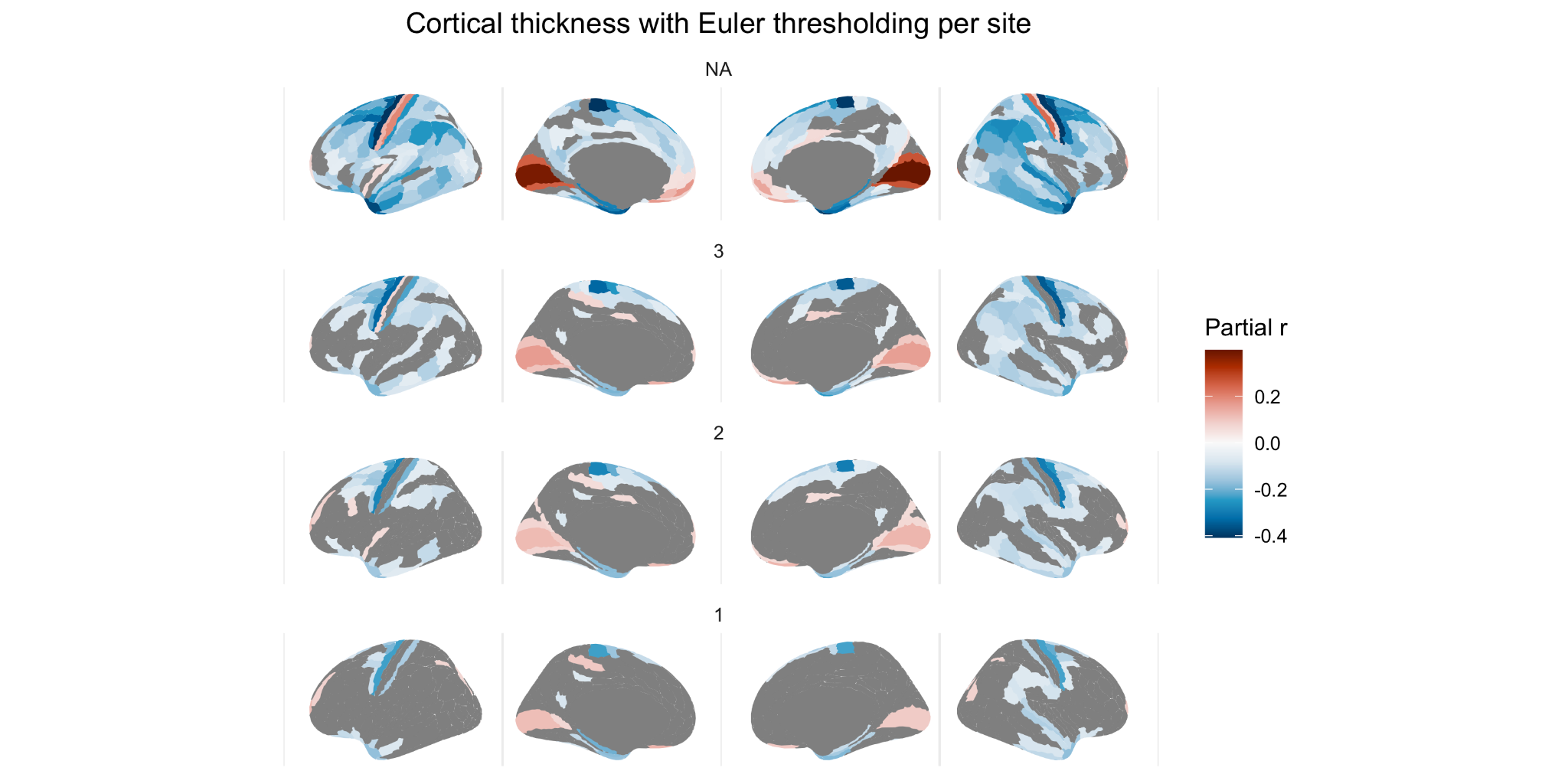

Fig S1.3. Relationship between CT and Euler at various Euler thresholds based on MADs applied per individual site (significant regions only).

##### 3.2 FSQC median split analysis (CT)

Our median split analysis essentially compared all those with perfect or near perfect scans and surface reconstructions, as rated by our FSQC, to those with minor or major errors (split below and above 1.1, with 1 being a rating of “good” on all 10 images). Here, we observed significant and widespread cortical thickness differences between groups, in similar areas to those in which we observed significant correlations between scan quality and CT. The group differences we observed were largely of greater CT in the “high” quality group relative to the “lower” quality group. This is consistent with our previous results suggesting apparent cortical thinning related to poor scan quality. Again, as in these previous analyses, the exceptions were in the medial occipital cortex, inferior medial prefrontal cortex, and (right) postcentral gyrus, where we observed significantly lower CT values in the “high quality” group.

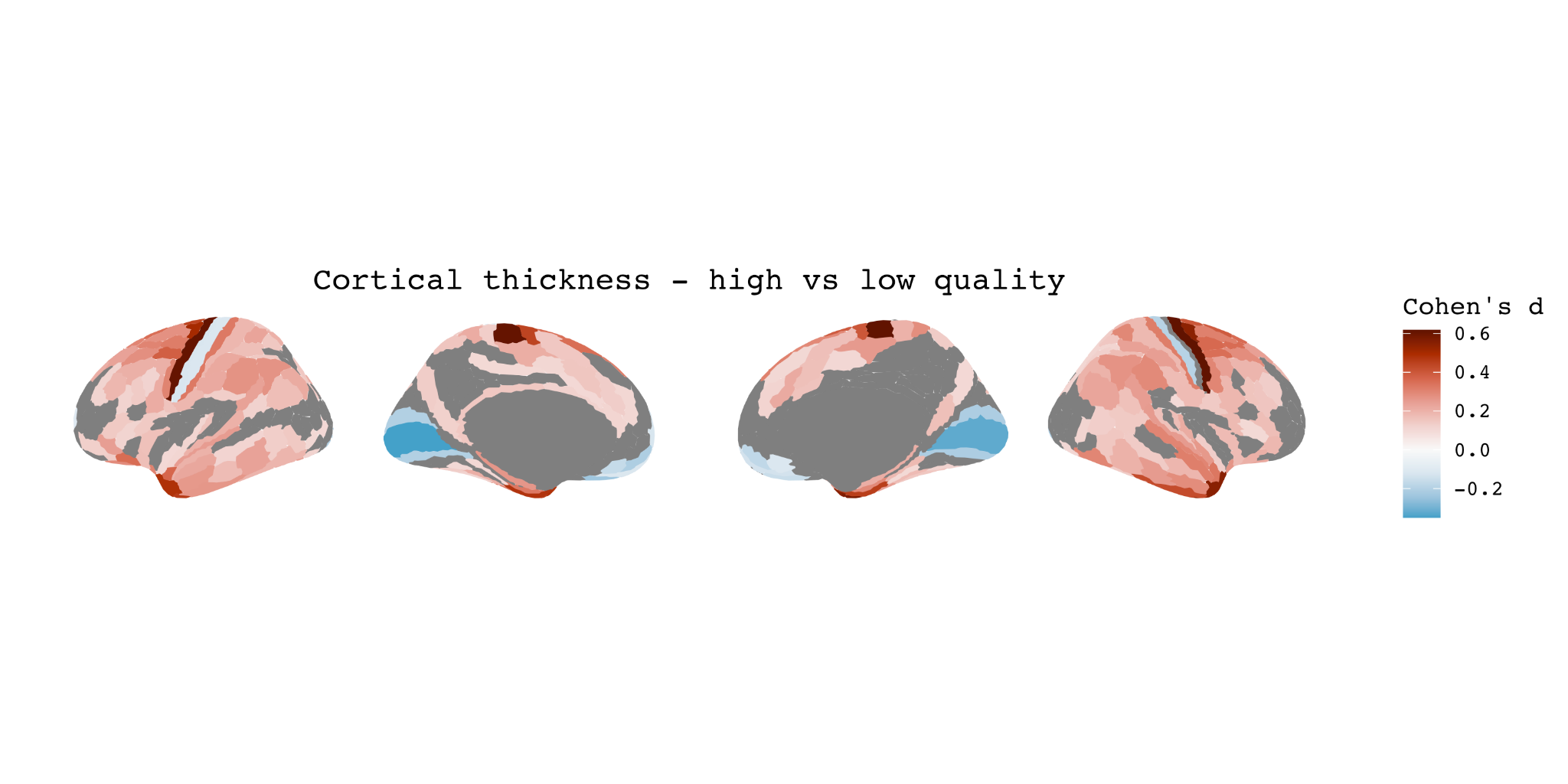

Fig S3.2 Median split analysis comparing scans with high (<1.1) vs low (>1.1) quality scans and outputs based on FSQC ratings.

##### 3.3 Percent exclusion analysis

In both analyses (exclusion based on whole sample and per site), results were further attenuated at every 5% threshold interval, such that after excluding 50% of the data, only a few significant associations remained. These included negative correlations in the precentral gyrus, inferior temporal cortex, and left postcentral gyrus, and positive correlations in the prefrontal cortex and left post central gyrus. However, in the whole sample analysis, more positive associations were observed in the frontal and parietal cortices at cut offs of 30-45%, whereas in the per site analysis, these effects were observed after 40-50% exclusion.

In these analyses, we also modelled the relationship between the percent of participants excluded, and the number of significant regions, as well as between the percent excluded and the effect size of the regions with the strongest effects. For both analyses, there was an initially strong negative relationship between percent excluded and number of significant regions. For the per-site analysis, the inflection point where the curve began to level off was 20; for the whole sample analysis, it was 15. The relationship between percent excluded and partial r values varied by region, but a similar pattern was seen to that of the number of significant regions .

**Participants excluded per site**

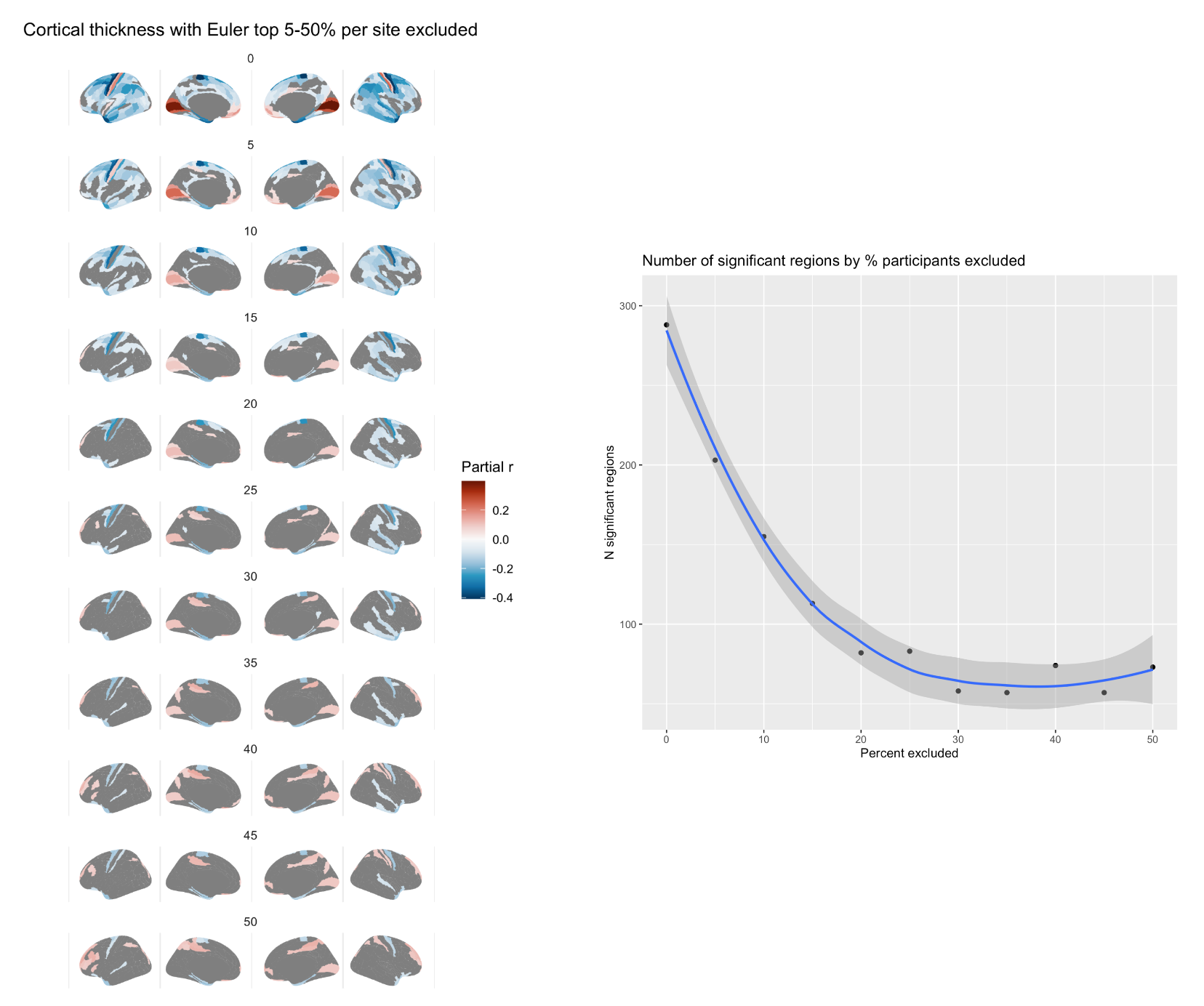

Fig S3.3.1. Effect of Euler number on CT after excluding top percent of participants per site, in intervals of 5% (left), and relationship between number of regions showing a significant relationship and percent of participants excluded (right).

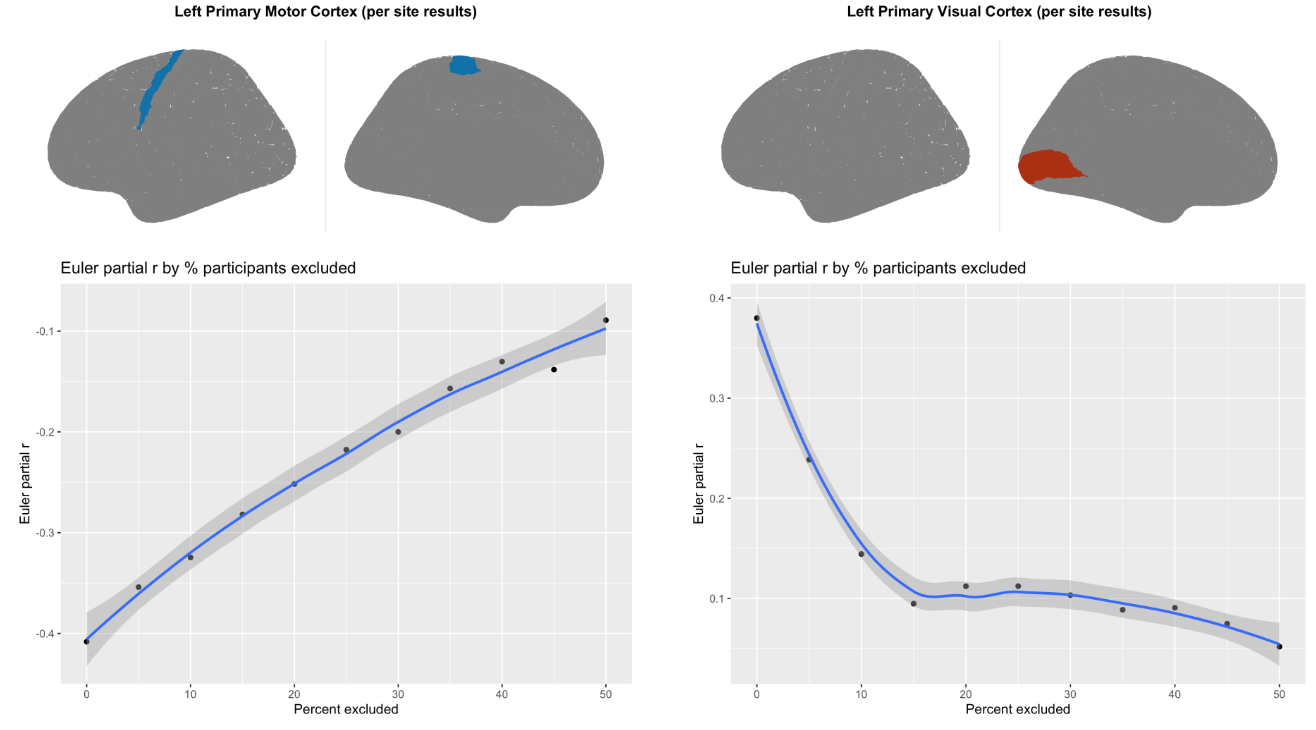

Fig S3.3.2. Relationship between correlation strength (partial r correlation for Euler number) and percent of participants excluded for two of the regions showing the strongest relationship: primary motor cortex (left) and primary visual cortex (right).

**Participants excluded from whole sample**

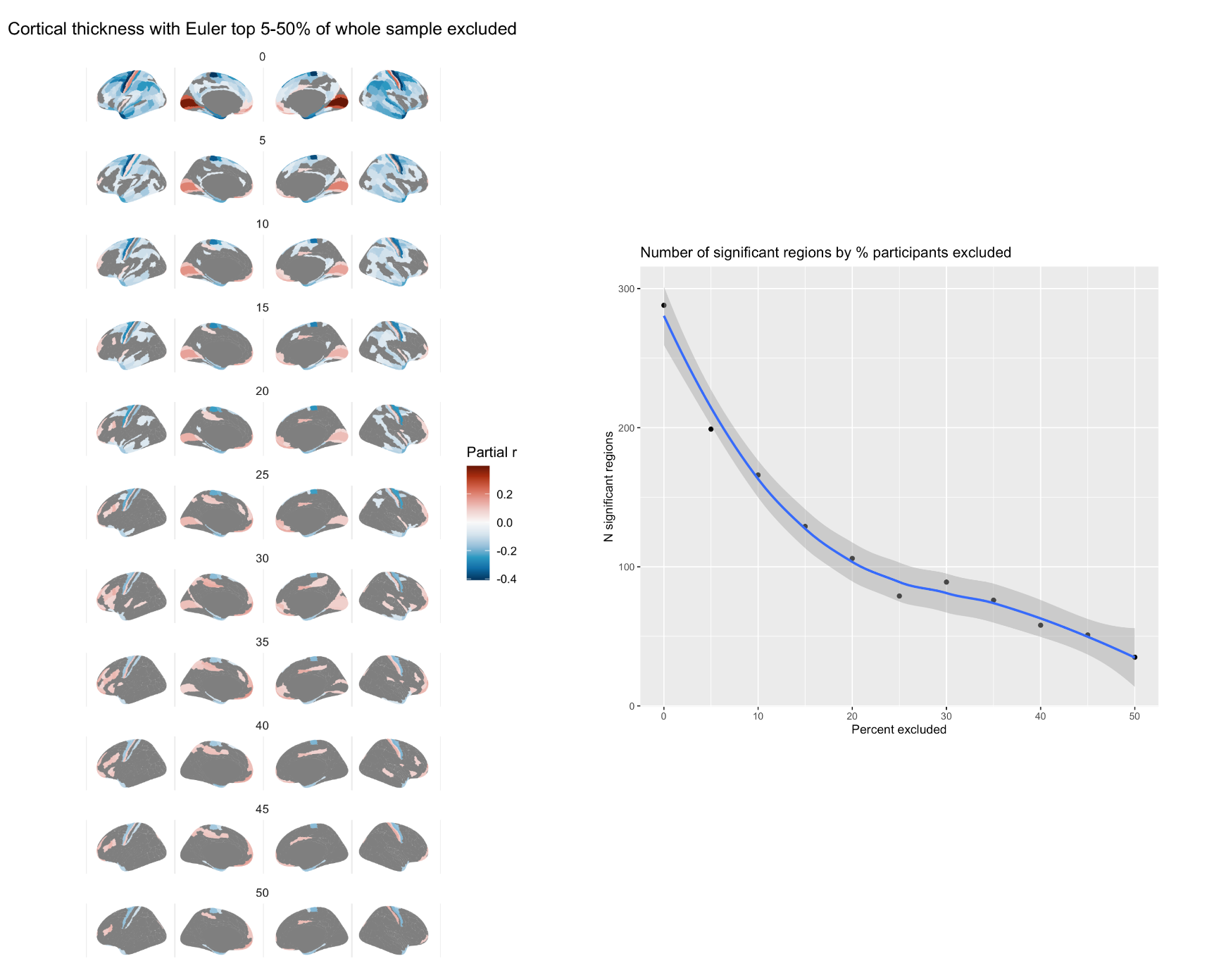

Fig S3.3.3 Fig Effect of Euler number on CT after excluding top percent of participants based on the whole sample, in intervals of 5% (left), and relationship between number of regions showing a significant relationship and percent of participants excluded (right).

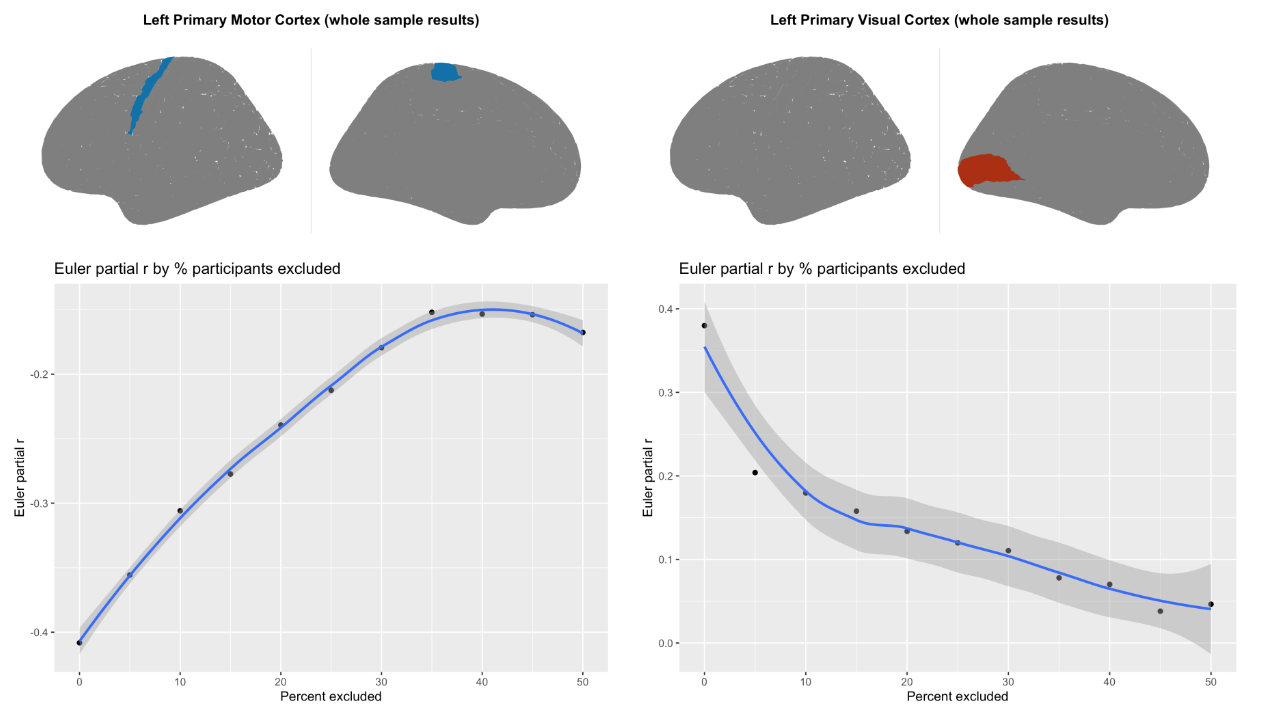

Fig S3.3.4. Relationship between correlation strength (partial r correlation for Euler number) and percent of participants excluded for two of the regions showing the strongest relationship: primary motor cortex (left) and primary visual cortex (right).

##### 3.4 SA and CV: FSQC thresholding

When thresholding by FSQC for SA and CV, there was a stark drop off in significant effects even when excluding only those above a score of 2.5, with very few significant regions remaining. Subthreshold maps (i.e. including non significant regions) showed a mix of positive and negative partial correlations after initial thresholding

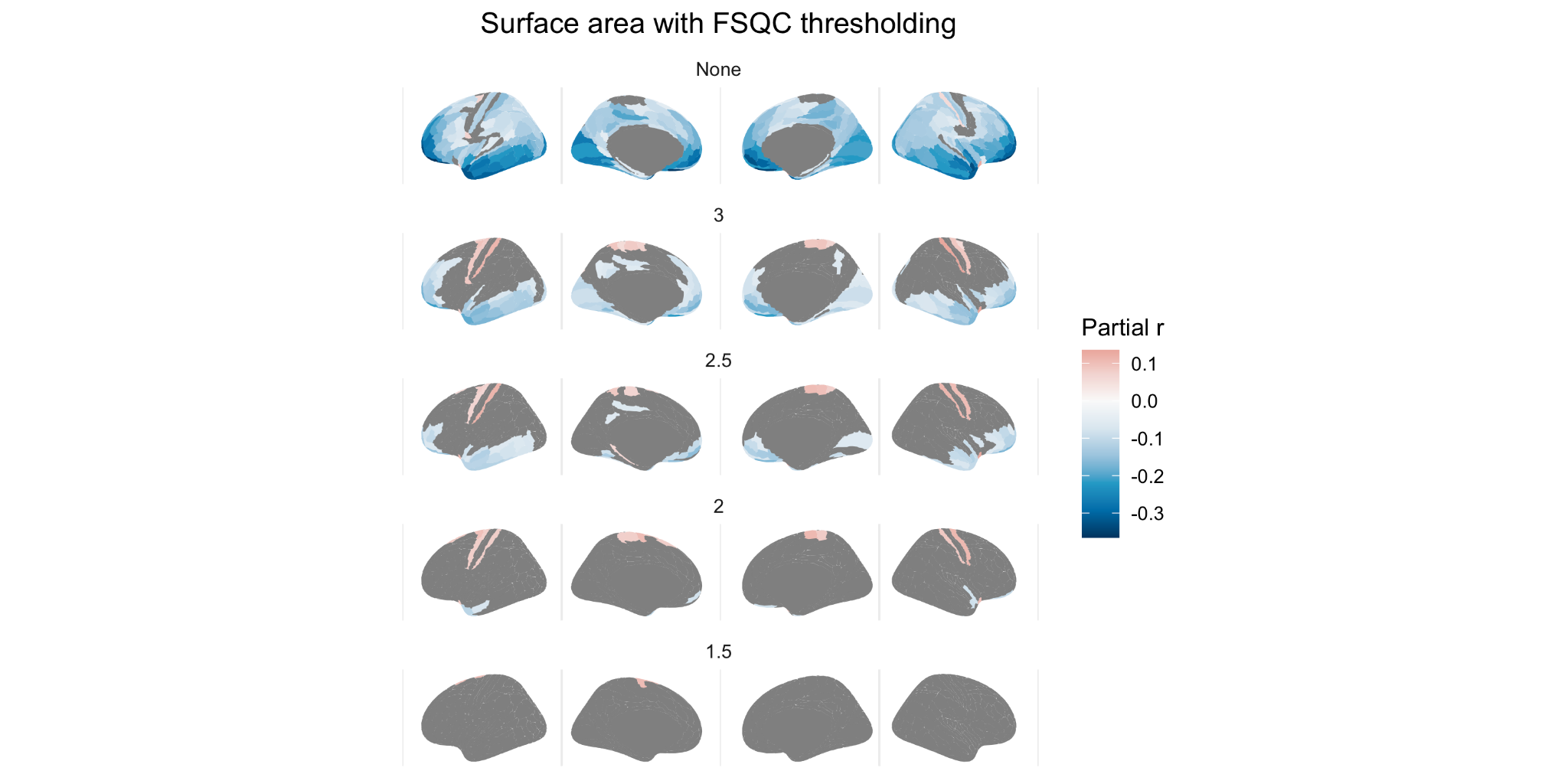

Fig S3.4.1 Relationship between SA and FSQC at various FSQC thresholds

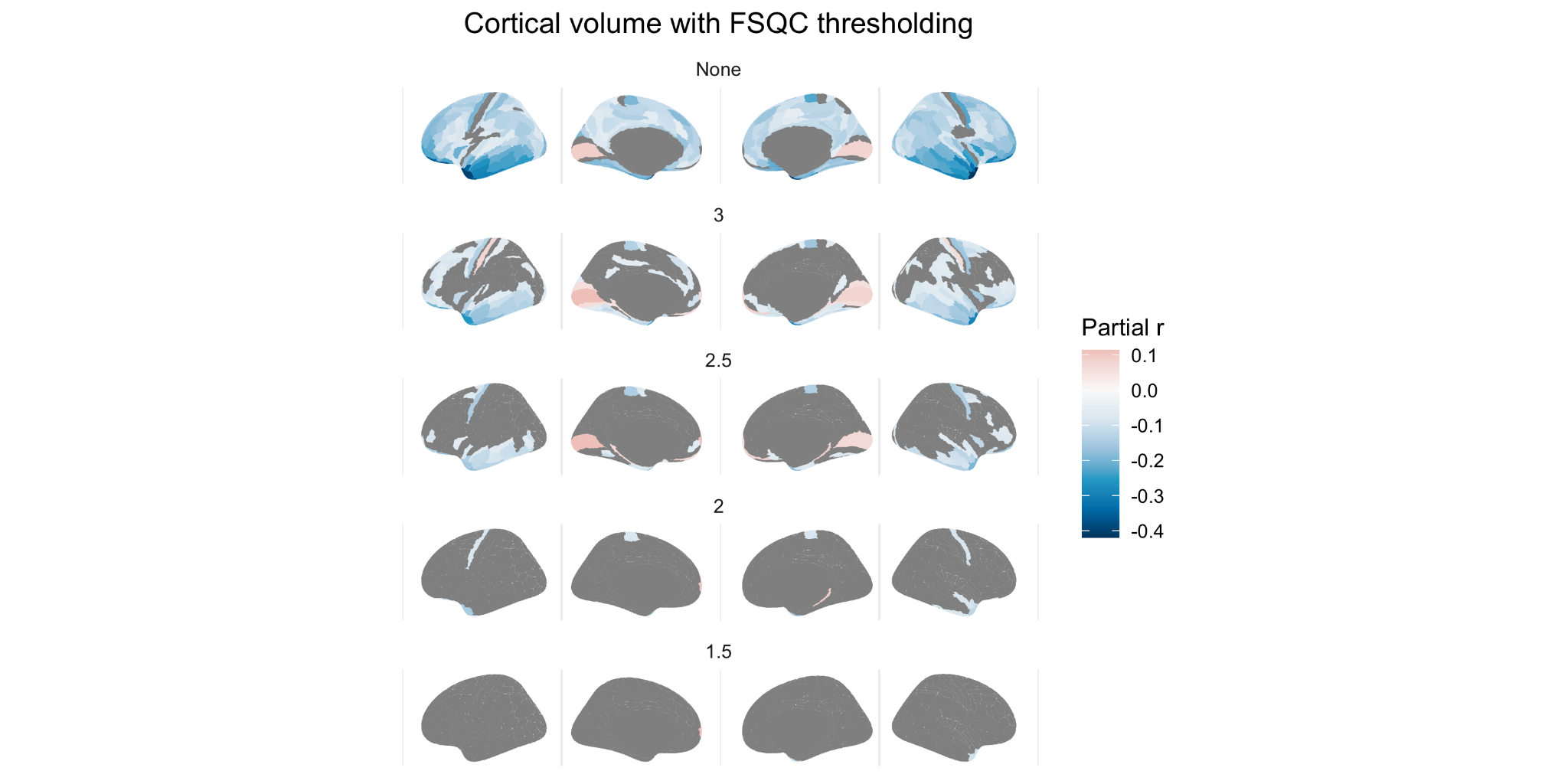

Fig S3.4.2 Relationship between CV and FSQC at various FSQC thresholds

##### 3.5 SA and CV: Euler thresholding

For SA and CV, when thresholding by Euler number, results largely demonstrated positive rather than negative correlations after even the least stringent threshold.

This appears to be largely due to different regions becoming significant, rather than a reversing of effect direction within regions. Positive results were not attenuated with lowering thresholds; in fact, they were observed in more widespread regions by the most stringent threshold. For SA, after thresholding at any of the MAD cut off points, largely positive significant correlations were observed, primarily in the superior frontal and temporal cortices, and parietal regions. For CV, the same is true, but primarily in prefrontal and parietal cortices. It should also be noted that when thresholding using FSQC, while very few regions remain significant after exclusions, spatial patterning is similar to those observed here.

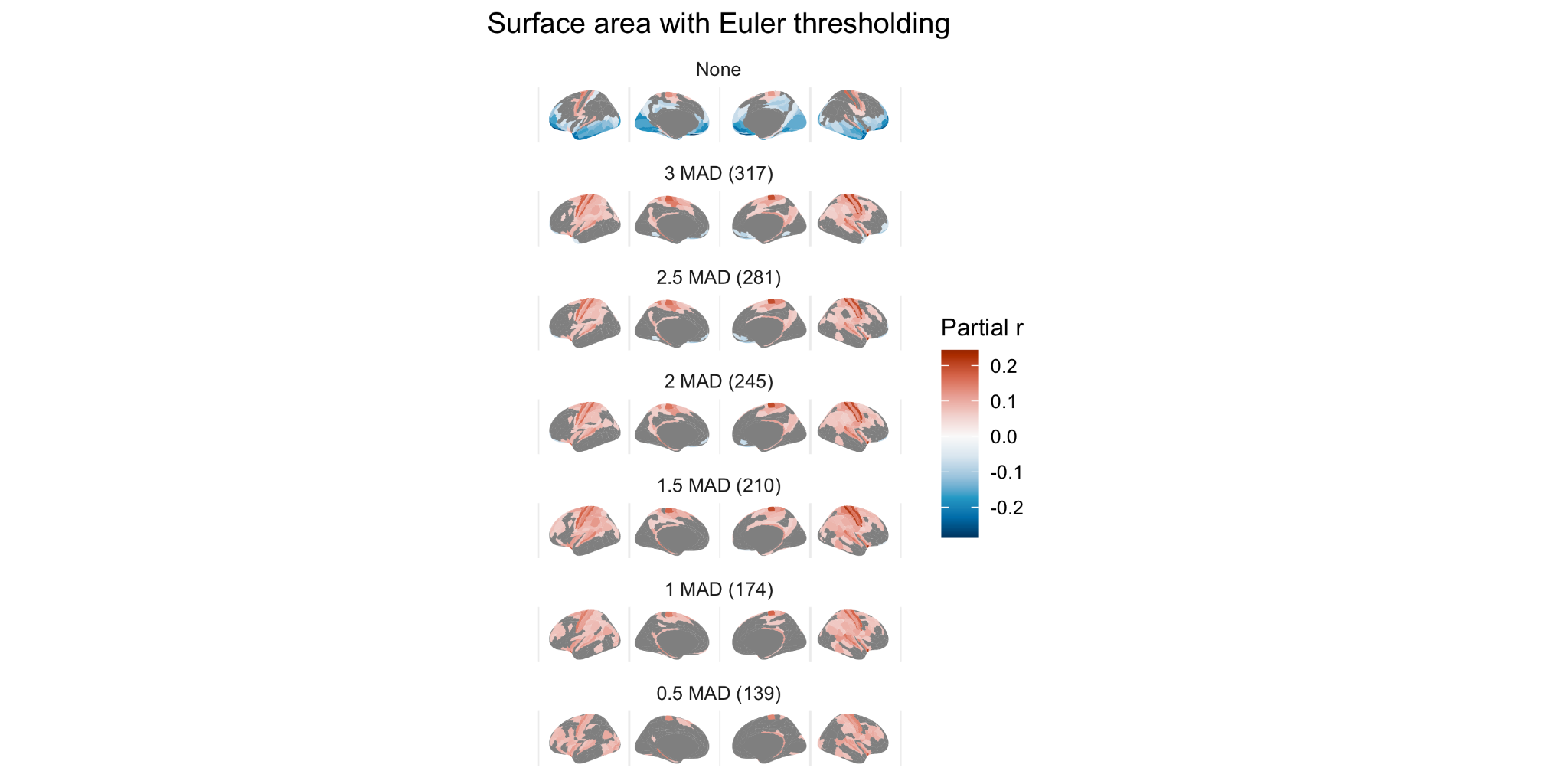

Fig S3.5.1 Relationship between SA and Euler at various Euler thresholds

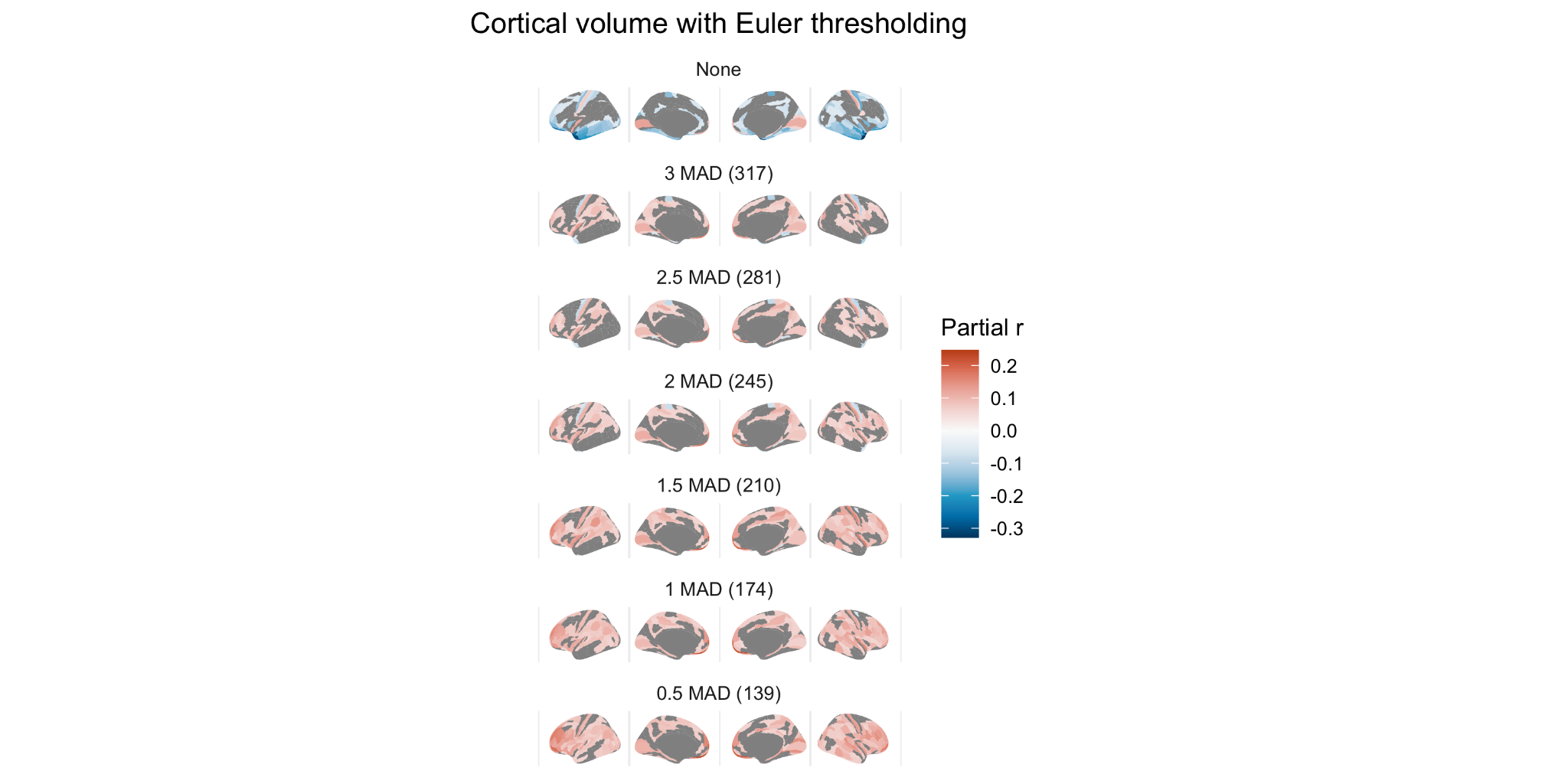

Fig S3.5.2 Relationship between CV and Euler at various Euler thresholds

#### 4. Diagnosis

##### 4.1 Diagnosis with cut off points

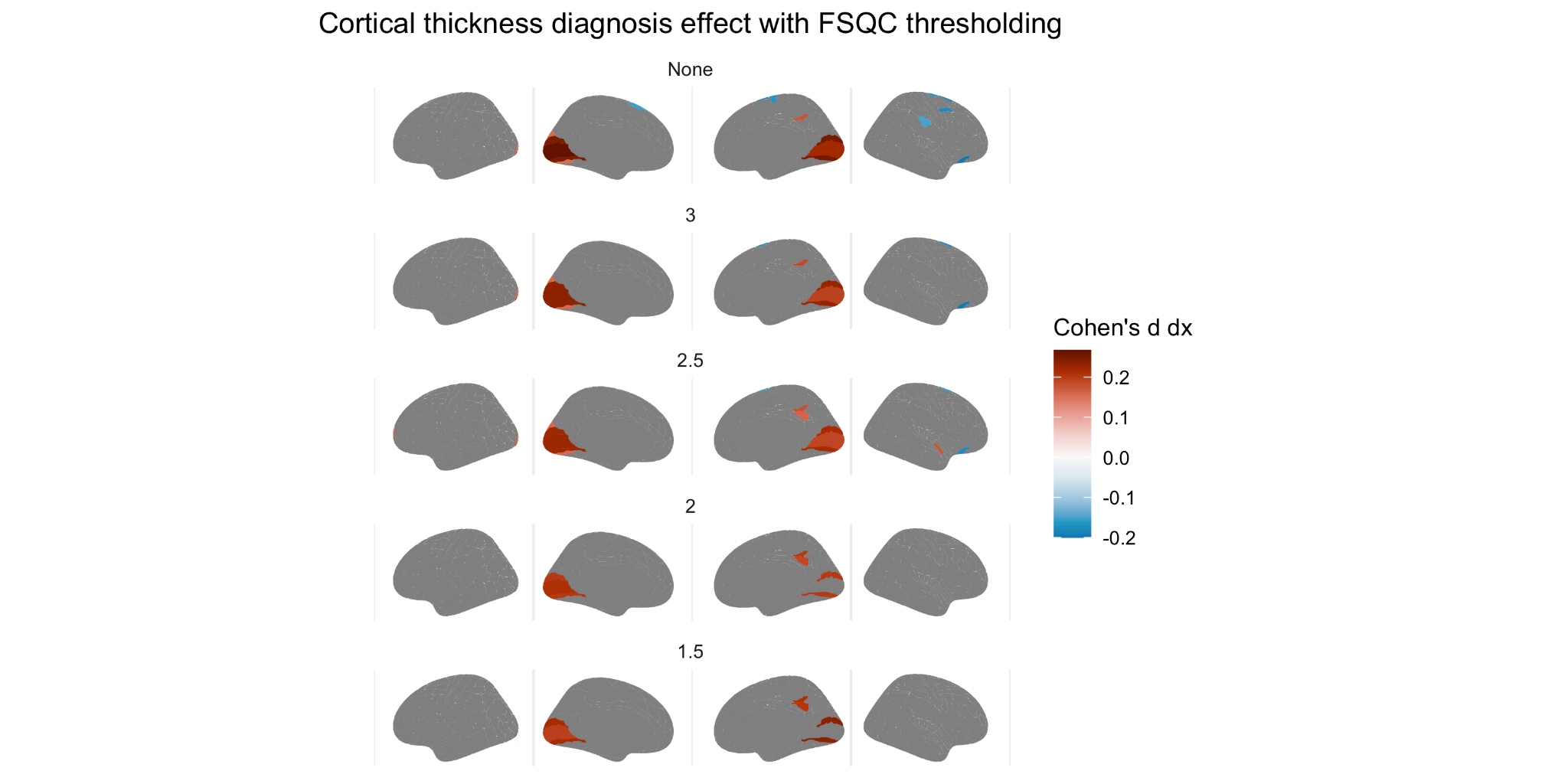

Fig S4.1.1. Effect of diagnosis (Cohen’s *d*) on cortical thickness at various thresholds of FSQC

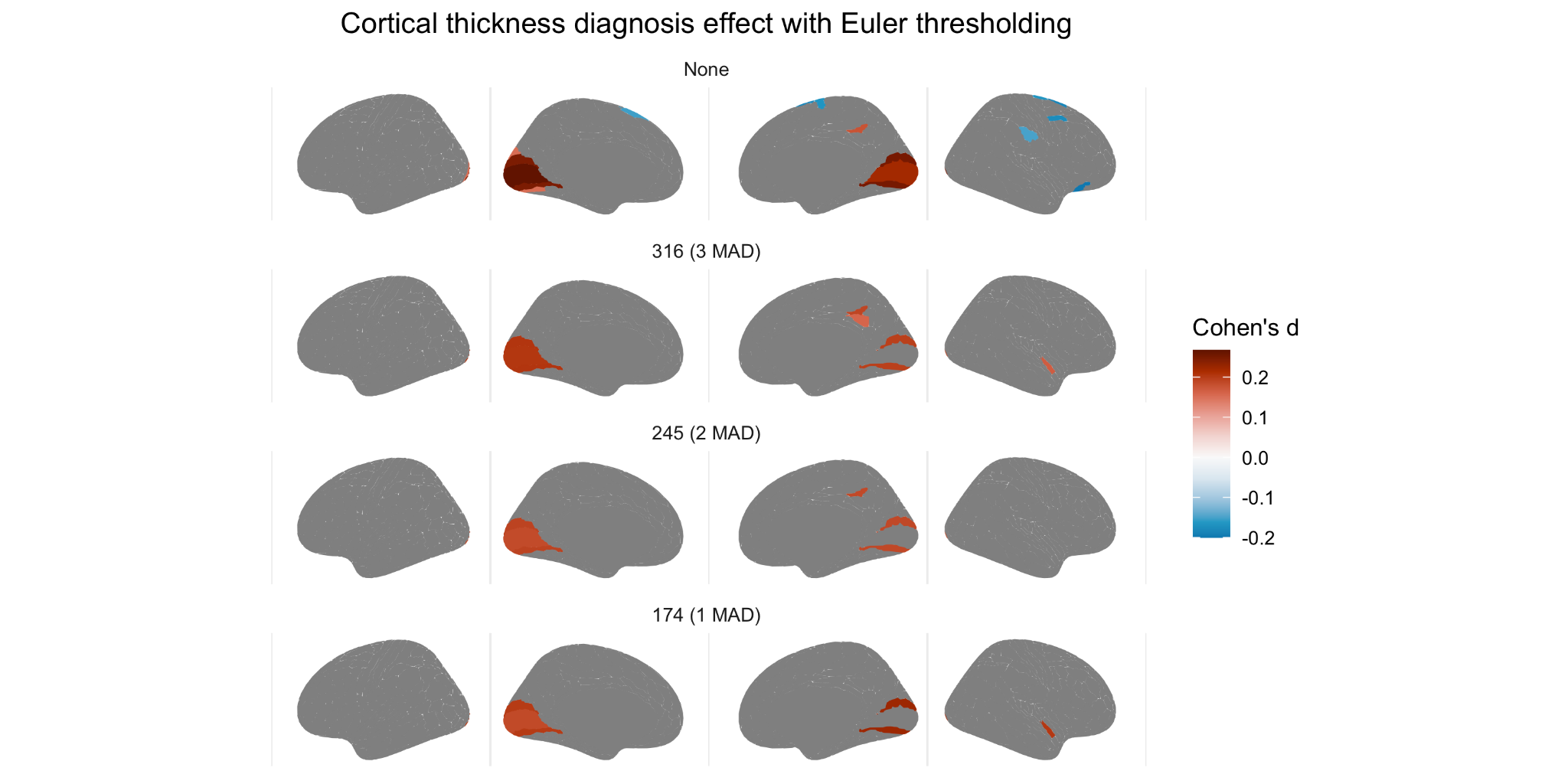

Fig S4.1.2. Effect of diagnosis (Cohen’s *d*) on cortical thickness at various thresholds of Euler

##### 4.2 Diagnosis FSQC threshold + Euler

We also combined these two approaches, by applying a threshold based on FSQC, and also controlling for Euler. In this analysis, results were very similar to the previous FSQC thresholding analysis, with the exception of additional significant regions (autism > controls) in the superior frontal and temporal regions.

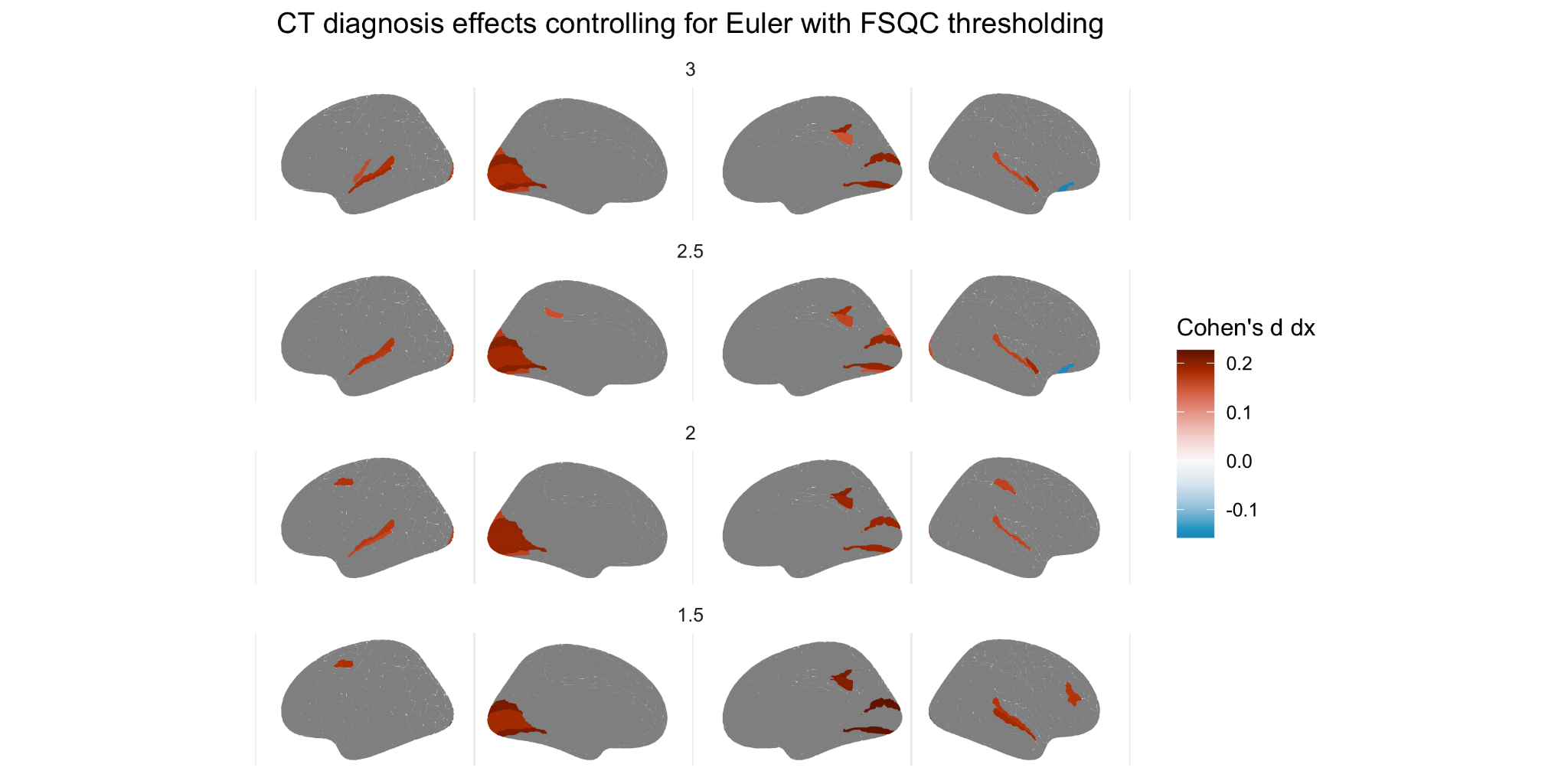

##### Fig S4.2. Effect of diagnosis on cortical thickness at various FSQC thresholds and controlling for Euler number.

##### 4.3 Diagnosis interaction

We also examined the interaction between diagnosis and scan quality for both Euler and FSQC. For FSQC, a significant interaction was observed in multiple frontal, temporal and parietal regions, in particular in the right hemisphere. For Euler index, very few significant regions were observed, including small regions in the medial frontal cortices and left posterior parietal cortex. To explore this further, we examined the association between quality and CT in autistic individuals and controls separately. In regions where we observed a significant interaction, correlations appeared to be stronger in autistic individuals than controls, though in the same direction.

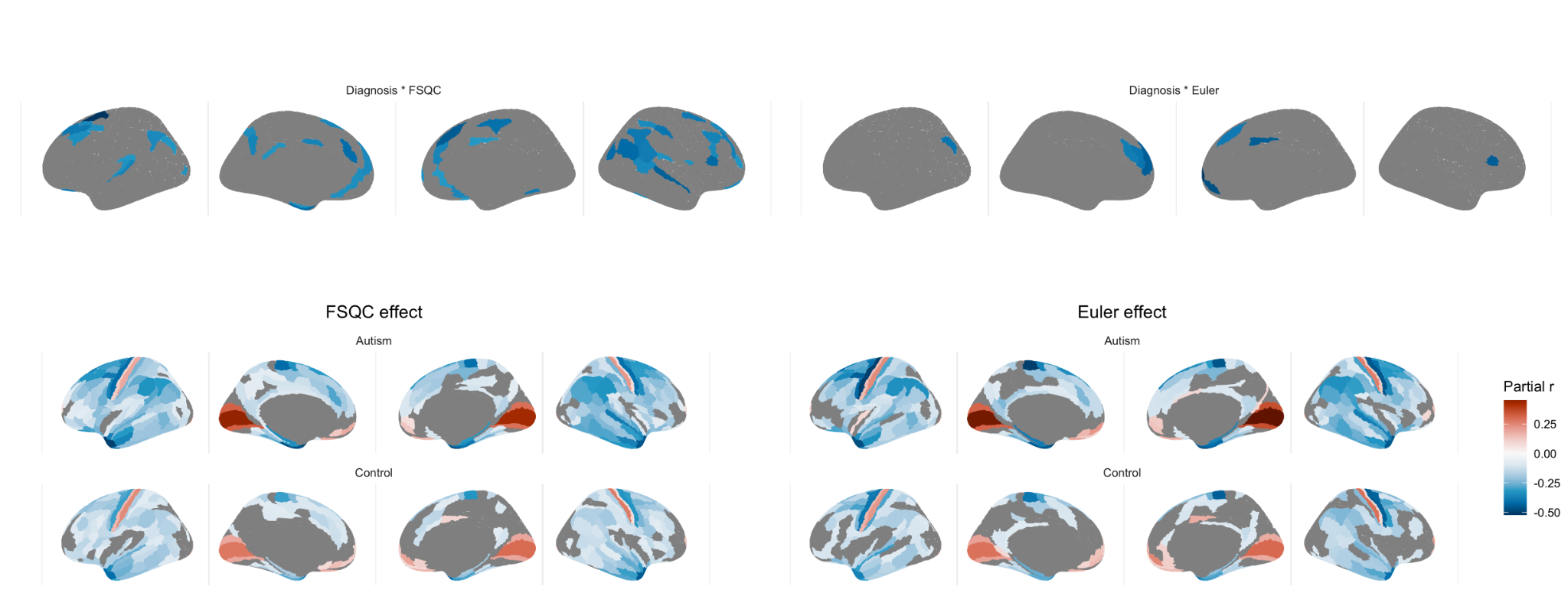

Fig 4.3 Interaction between diagnosis and FSQC (top right) and diagnosis and Euler number (top left), and effect of FSQC (bottom right) and Euler number (bottom right) in the autism and control groups, respectively.

##### 4.4 Diagnosis SA and CV

Only very minimal group differences in SA and CV were observed, both with and without accounting for scan quality. For SA, without accounting for quality, lower SA in autistic individuals compared to controls was observed in right V1 only. After controlling for either FSQC or Euler, no significant results remained. For CV, no significant associations were observed before controlling for quality; however, after controlling for either FSQC or Euler, a significant effect was observed in the superior temporal gyrus, in which autistic individuals had greater cortical volume relative to controls.

**SA:**

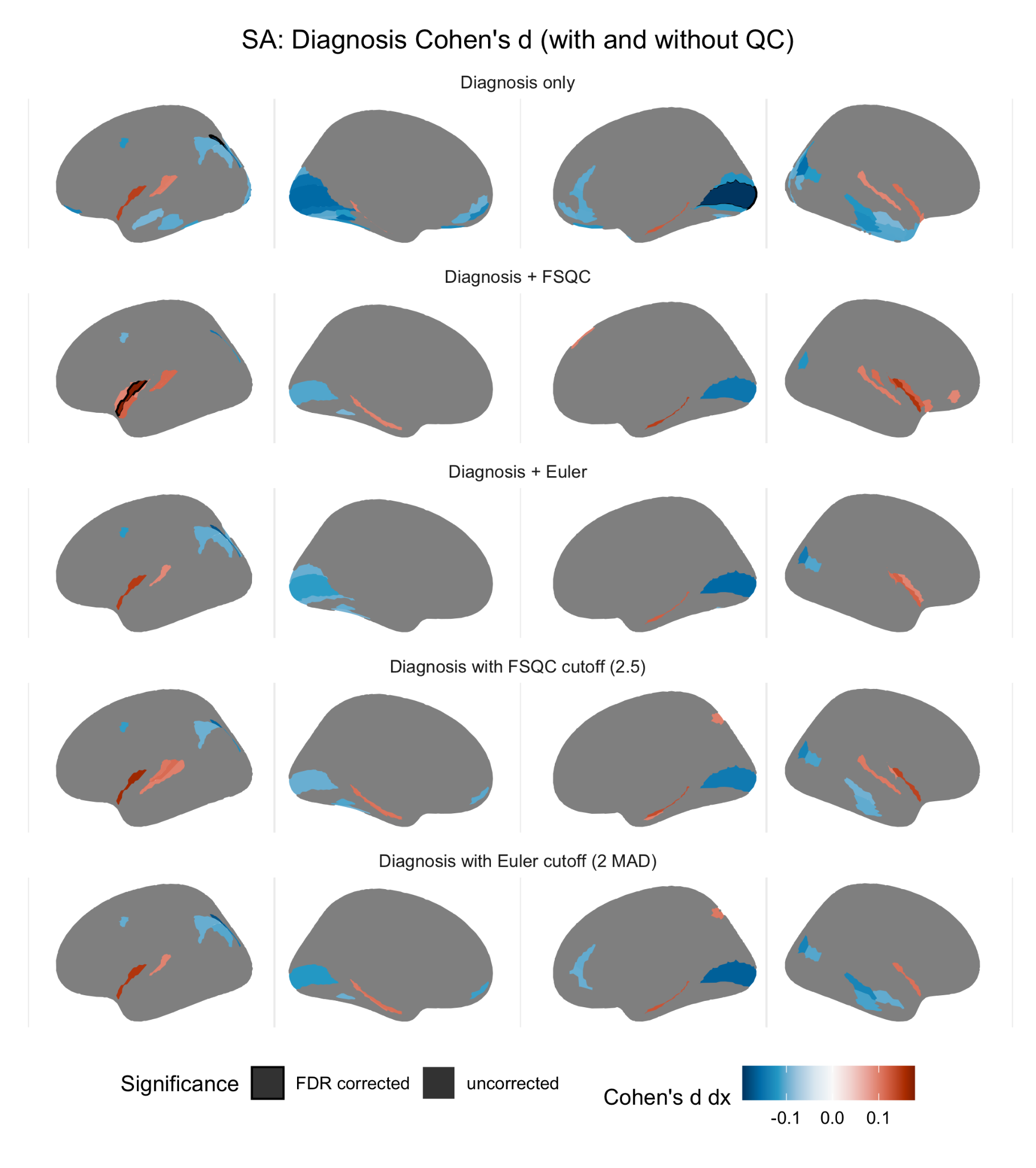

Fig S4.4.1. Impact of autism diagnosis on cortical surface area (Cohen’s *d*) without accounting for scan quality (A), when controlling for FSQC (B) or Euler (C), and thresholding by FSQC (D) and Euler (E). Significant regions passing 5% FDR are shown with a black border; other regions are subthreshold (not surviving FDR) differences.

**CV:**

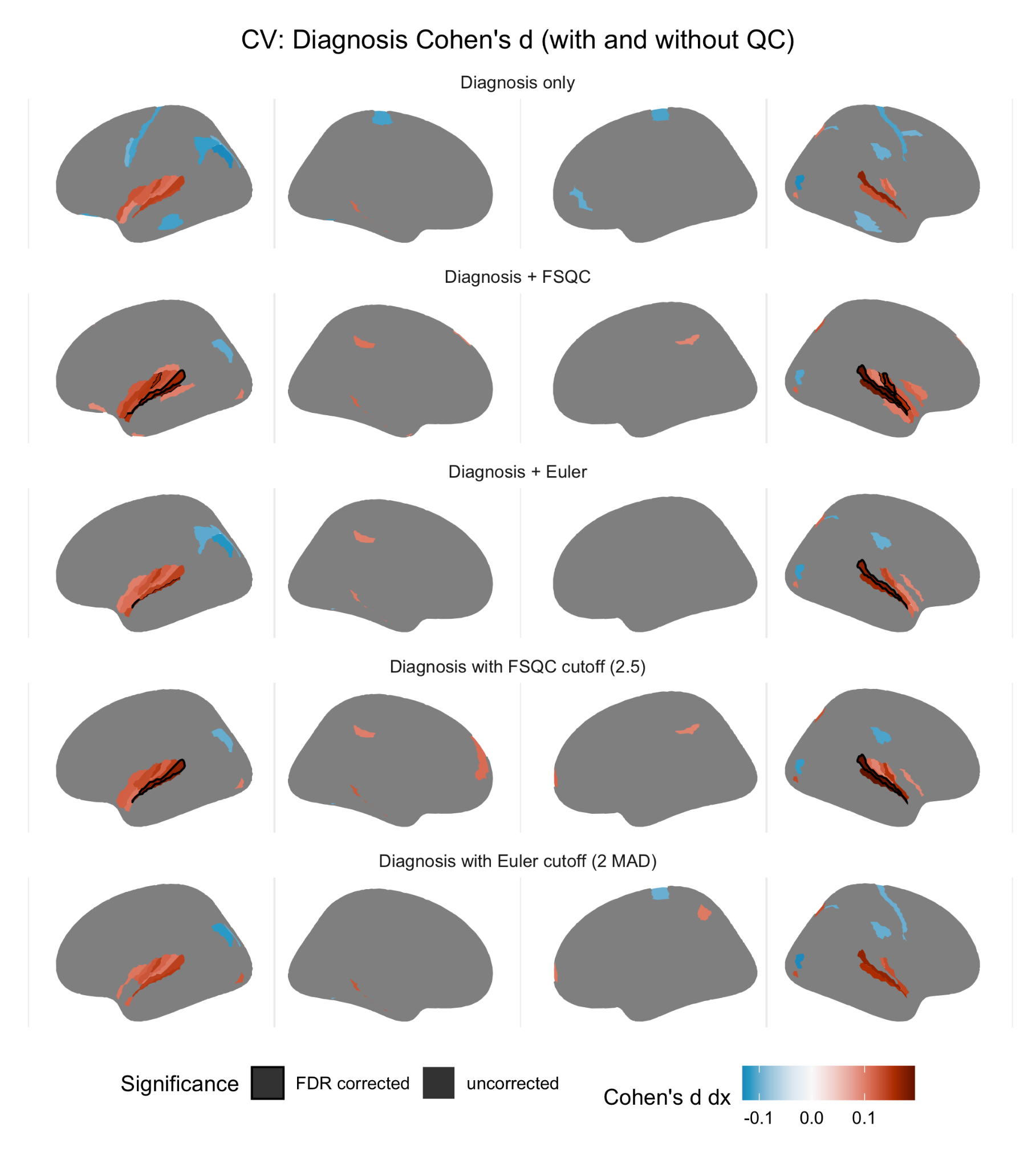

Fig S4.4.2. Impact of autism diagnosis on cortical volume (Cohen’s *d*) without accounting for scan quality (A), when controlling for FSQC (B) or Euler (C), and thresholding by FSQC (D) and Euler (E). Significant regions passing 5% FDR are shown with a black border; other regions are subthreshold (not surviving FDR) differences.

Hoffman, G. E., & Schadt, E. E. (2016). variancePartition: interpreting drivers of variation in complex gene expression studies. *BMC Bioinformatics*, *17*(1), 483.
